# Latent Class Trajectory Phenotypes of Longitudinal Visit and Follow-up Patterns Among Patients with Hypertension

**DOI:** 10.64898/2026.07.22.26358744

**Authors:** Haoxin Chen, Jiancheng Ye

## Abstract

**Objective:** To identify latent phenotypes of observed electronic health record (EHR) contact among patients with hypertension, evaluate their reproducibility across temporal resolutions and sensitivity to administrative censoring, and examine associations with treatment documentation and blood pressure (BP) control after adjustment for comorbidity burden and measurement opportunity.

**Methods:** We conducted a retrospective cohort study of patients with hypertension and at least two recorded visits (N = 26,710). Latent class trajectory modelling of repeated binary visit indicators within a Bernoulli finite mixture framework was performed using 1- month and 3-month intervals over 36 months. Models were selected using the Bayesian Information Criterion and GRoLTS-recommended metrics. Multivariable linear regression examined associations between trajectory phenotypes and treatment documentation and BP control after adjustment for age, sex, race, baseline BP, and comorbidity burden. BP outcomes were also compared at 12, 24, and 36 months.

**Results:** Four observed EHR contact trajectory groups were identified at both temporal resolutions (1-month: relative entropy 0.821, minimum APP 0.770, minimum OCC 6.49; 3-month: relative entropy 0.771, minimum APP 0.778, minimum OCC 5.39), with excellent bootstrap reproducibility (mean ARI 0.966–0.969). The four-group structure was replicated in the ≥ 24-month subgroup, although this represented only 20.8% of the cohort. Overall, 79.2% of patients had <24 months of follow-up, with censoring concentrated in the lowest-contact groups, indicating that these trajectories reflect observed EHR contact under variable administrative observation rather than patient disengagement. After adjustment, higher-contact groups had consistently higher treatment documentation rates than the low observed follow-up group, whereas differences in BP control were small and inconsistent. The highest-contact group did not achieve the lowest BP at any fixed time point despite the largest first-to-last BP reduction, demonstrating bias from differential observation window length.

**Conclusion:** Four internally reproducible phenotypes of observed EHR contact were identified, but their trajectories were substantially influenced by administrative censoring and lack external validation. Contact phenotype was associated with treatment documentation but only weakly with BP control after adjustment.

## INTRODUCTION

Hypertension is a leading modifiable risk factor for cardiovascular morbidity and mortality, affecting an estimated 1.28 billion adults worldwide in 2019.[1] Effective long-term blood pressure (BP) control depends not only on timely diagnosis and pharmacotherapy but also on sustained retention in care through regular follow-up, BP monitoring, and treatment adjustment.[2] Despite evidence-based guidelines, BP control remains suboptimal in routine practice, partly because many patients discontinue follow-up within the first year after diagnosis.[3, 4]

Retention in care, engagement, and medication adherence are related but distinct constructs: retention in care (continued presence in the healthcare system as evidenced by visit attendance), engagement (active and purposeful participation in care processes), and adherence (medication-taking behavior).[5] Electronic health record (EHR) data capture visit attendance and therefore measure retention, but cannot directly assess engagement or adherence. Distinguishing these constructs is essential to avoid overstating the clinical implications of EHR-based analyses.[5] Although trajectory analyses in hypertension have predominantly examined medication adherence using pharmacy refill measures and linked adherence patterns to cardiovascular outcomes, [6] trajectory-based studies of visit retention in hypertension care remain scarce, representing an important evidence gap.[7]

Longitudinal follow-up patterns in hypertension are inherently heterogeneous. While some patients maintain regular follow-up, others disengage early or exhibit intermittent attendance.[8] Conventional summary measures, such as total visit counts or follow-up duration, assume population homogeneity and may obscure clinically meaningful subgroups. Finite mixture models address this limitation by identifying latent groups with similar longitudinal patterns. Group-based trajectory modelling (GBTM), originally developed by Nagin,[9] has been widely applied in pharmacoepidemiology, behavioral research,[10] and chronic disease studies, including glycemic control.[11] In this study, we applied latent class trajectory modelling of repeated binary visit indicators within a Bernoulli finite mixture framework. Unlike classical GBTM, which models polynomial trajectories, this approach estimates class-specific attendance probabilities at each time point, providing greater flexibility for irregular, non-monotonic EHR follow-up patterns. Compared with growth mixture models, it assumes no within-class variability around a mean trajectory, and compared with sequence analysis, it provides probabilistic class membership and likelihood-based model selection using the Bayesian Information Criterion (BIC). Despite these advantages, its application to visit-based EHR follow-up in hypertension has been limited.[12]

We applied this approach to identify latent patterns of observed EHR contact among patients with hypertension during the first 36 months after the index visit. Visit attendance was modelled as a binary outcome at both 1-month and 3-month intervals to evaluate follow-up at different temporal resolutions. Baseline demographic and clinical characteristics were compared across trajectory groups, and associations with treatment documentation and BP control were examined. Consistent with the ABC taxonomy,[5] all inferences were restricted to observed visit-based retention. By characterizing distinct EHRs contact trajectories, this study aims to improve understanding of heterogeneity in hypertension follow-up and support identification of patients at risk of early disengagement from observed care. Clinical associations are interpreted cautiously because observed contact is influenced by administrative observation windows and opportunities for measurement.

## METHODS

### Study design

This study used deidentified EHR data from primary care practices participating in the EvidenceNOW quality-improvement program, which supports small-to-medium primary care practices across multiple U.S. regions in delivering evidence-based cardiovascular preventive care.[13] The database comprised four datasets: (1) Diagnosis (patient level), including encounter dates and ICD/SNOMED CT codes for hypertension and related comorbidities; (2) Demographics (patient level), including age, sex, race, ethnicity, and language; (3) Vitals (visit level), including blood pressure (BP), body mass index, height, and weight; and (4) Medications (visit level), including medication names, codes, and prescription start and end dates. Data were contributed by multiple primary care sites. Because follow-up was censored at a fixed extraction date, observable follow-up varied according to the timing of each patient’s index visit.

### Measures

The hypertension cohort was identified using ICD diagnosis codes supplemented by keyword searches of diagnosis records. Records with missing patient identifiers were excluded. The index visit was defined as the first hypertension-related encounter, and follow-up was measured from this date. A visit was defined as any calendar day with at least one hypertension-related encounter, with multiple encounters on the same day treated as a single visit. Patients with fewer than two visits were excluded to permit longitudinal trajectory estimation.

BP control was defined as systolic BP <140 mmHg and diastolic BP <90 mmHg; the guideline-recommended threshold of <130/80 mmHg is addressed in the Limitations. Treatment status at each visit was defined by whether the visit date fell within a recorded antihypertensive prescription period based on prescription start and end dates. Patient-level treatment documentation and BP control rates were calculated as the proportions of visits with documented antihypertensive treatment and controlled BP, respectively. The treatment documentation rate reflects recorded prescription coverage at visits rather than medication adherence.

### Descriptive analysis

Descriptive analyses characterized longitudinal patterns of observed EHR contact, consistent with the ABC taxonomy of adherence-related constructs[5]. The proportion of patients with at least one recorded encounter was estimated at 6, 12, 18, 24, 30, and 36 months after the index visit. Total visit count and follow-up duration were summarized using medians and interquartile ranges. Among patients who experienced prolonged interruptions in care, the time to the first inter-visit gap exceeding 6, 12, and 18 months was estimated. BP control rates were assessed at 12, 24, and 36 months using the closest BP measurement within a ±3-month window. The number and proportion of patients with available BP measurements were reported at each time point because measurement availability depended on observed contact and was unlikely to be random.

### Latent Class Trajectory Modeling of Binary Visit Indicators

Latent class trajectory modelling of repeated binary visit indicators was used to identify subgroups of patients with distinct longitudinal patterns of observed EHR contact. The first 36 months after the index visit were divided into either 36 one-month intervals or 12 three-month intervals. For each interval, the outcome was a binary indicator of visit attendance, defined as the presence of at least one recorded hypertension-related encounter.

### Models

Latent class trajectory models were estimated within a Bernoulli finite mixture framework without random effects using StepMix v3.0.0.**[14]** Unlike classical group-based trajectory modelling (GBTM), which specifies polynomial trajectories within each class, this approach estimated class-specific probabilities of visit attendance independently at each time interval, allowing flexible, non-parametric trajectory shapes. Let *y*_*it*_ ∈ {0,1} denote visit attendance for individual *i* at time interval *t*, and *Z*_*i*_ ∈ {1, …, *K*} latent class membership. The model assumes that the distribution of the observed longitudinal outcomes is governed by a mixture of K latent groups:

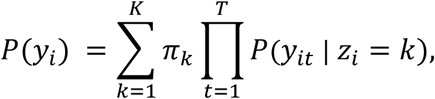

where π_*k*_ represents the class membership probability for group *k*, with 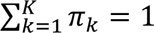. Conditional on class membership, visit outcomes across time intervals are assumed independent and Bernoulli distributed:

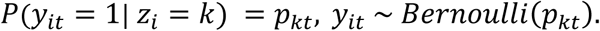

Model parameters were estimated via the expectation-maximization algorithm, using multiple random initializations and retaining the solution with the highest log-likelihood to reduce sensitivity to local maxima. Each individual was assigned to the trajectory group with the highest posterior probability (modal assignment):

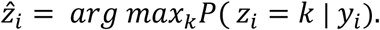

Within each interval, a value of 1 was assigned if at least one encounter was recorded and 0 otherwise, including intervals after a patient’s last recorded encounter. Because post-last-contact intervals coded as 0 cannot be distinguished from confirmed non-attendance, these values represent unverifiable absence rather than confirmed non-attendance. An alternative implementation treating post-last-contact intervals as missing via Full Information Maximum Likelihood was evaluated but produced severely degraded model fit across all K values in both resolutions (minimum OCC 0.84–1.84; relative entropy 0.256–0.564), and was therefore not adopted. This coding cannot distinguish administrative censoring from genuine non-attendance and is the principal interpretive constraint on the trajectory shapes. The resulting trajectory groups represent observed EHR contact patterns under variable observation windows and should not be interpreted as confirmed behavioral disengagement.

Models with *K* = 1 to *K* = 6 latent groups were estimated. Model selection was guided by the Bayesian Information Criterion (BIC) and clinical interpretability of the resulting trajectory shapes. To comply with the GRoLTS checklist for group-based trajectory model reporting [11] (**Supplementary Table S3**), the following model quality metrics were computed for each candidate model. Average posterior probability (APP) per group was calculated as the mean of each individual’s posterior probability of belonging to their assigned class; values > 0.70 are considered acceptable and > 0.80 good. Relative entropy was calculated as 1 − *H*^ˉ^/*log* (*K*), where *H*^ˉ^ is the mean per-individual Shannon entropy of the posterior probability vector; values > 0.80 indicate good class separation. Odds of correct classification (OCC) per group were derived from APP and the group’s estimated proportion 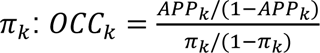; values > 5 indicate good separation. A full GRoLTS model fit table (BIC, AIC, log-likelihood, relative entropy, minimum APP, and minimum OCC across groups) is reported for both 1-month and 3-month interval models.

Class-assignment stability was evaluated using 200 bootstrap resamples. In each iteration, the model was refitted and agreement with the original classification was quantified using the Adjusted Rand Index (ARI). Bootstrap-derived 95% confidence intervals were also used for trajectory plots. To assess the impact of variable observation windows, the distribution of follow-up duration was summarized by trajectory group, and a prespecified sensitivity analysis restricted to patients with ≥24 months of observed follow-up was performed. Concordance between the 1-month and 3-month models was assessed using cross-tabulation, ARI, and Cohen’s κ.

### Group comparisons

Baseline demographic and clinical characteristics were compared across trajectory groups, including age, sex, race/ethnicity, comorbidities (diabetes, heart failure, stroke, myocardial infarction, and chronic kidney disease), baseline BP, treatment documentation, and BP control. Continuous variables were summarized as means (SD) or medians (IQR) and compared using Kruskal–Wallis tests. Categorical variables were summarized as counts (percentages) and compared using χ² or Fisher’s exact tests.

Between-group BP differences were quantified using standardized mean differences (SMDs) with 95% bootstrap confidence intervals (300 resamples), using the Low Observed Follow-up group as the reference. Within-patient changes in systolic and diastolic BP were assessed using Wilcoxon signed-rank tests, while changes in treatment documentation and BP control were evaluated using McNemar tests. BP at 12, 24, and 36 months was compared using the closest measurement within a ±3-month window.

Multivariable linear regression models were used to examine associations between trajectory group and patient-level treatment documentation and BP control rates, adjusting for age, sex, race/ethnicity, baseline systolic BP, diabetes, heart failure, stroke, and chronic kidney disease. Posterior probability-weighted analyses were performed as a sensitivity analysis. Clinical outcomes were visualized using boxplots and bar plots with 95% confidence intervals.

### Missing Data

Patients with missing identifiers were excluded during cohort construction. Age and treatment documentation were complete. BP data were unavailable for 1,147 patients (4.3%), who were excluded from BP control analyses. Race/ethnicity was available for all patients; however, 53% were recorded as unknown or unclassifiable and were retained as a separate category. Accordingly, race/ethnicity comparisons are descriptive.

### Study Approval

This study used de-identified data from the EvidenceNOW quality improvement program. The study was approved by the Northwestern University Institutional Review Board, which waived the requirement for informed consent.

## RESULTS

### Cohort Selection

The quality improvement program included 609,318 patients across participating primary care sites. Among these, 40,747 (6.7%) had a hypertension-related diagnosis identified using ICD and SNOMED CT codes supplemented by keyword searches of diagnosis records. Records with missing patient identifiers were excluded before linkage across diagnosis, demographics, vitals, and medication datasets.

Patients with only one recorded hypertension-related visit were excluded, yielding a final cohort of 26,710 patients (65.5% of those with hypertension; 4.4% of the overall program population) (**Figure 1**). This restriction ensured sufficient longitudinal data for trajectory modelling and excluded isolated encounters unlikely to represent ongoing hypertension management.

**Figure 1.**
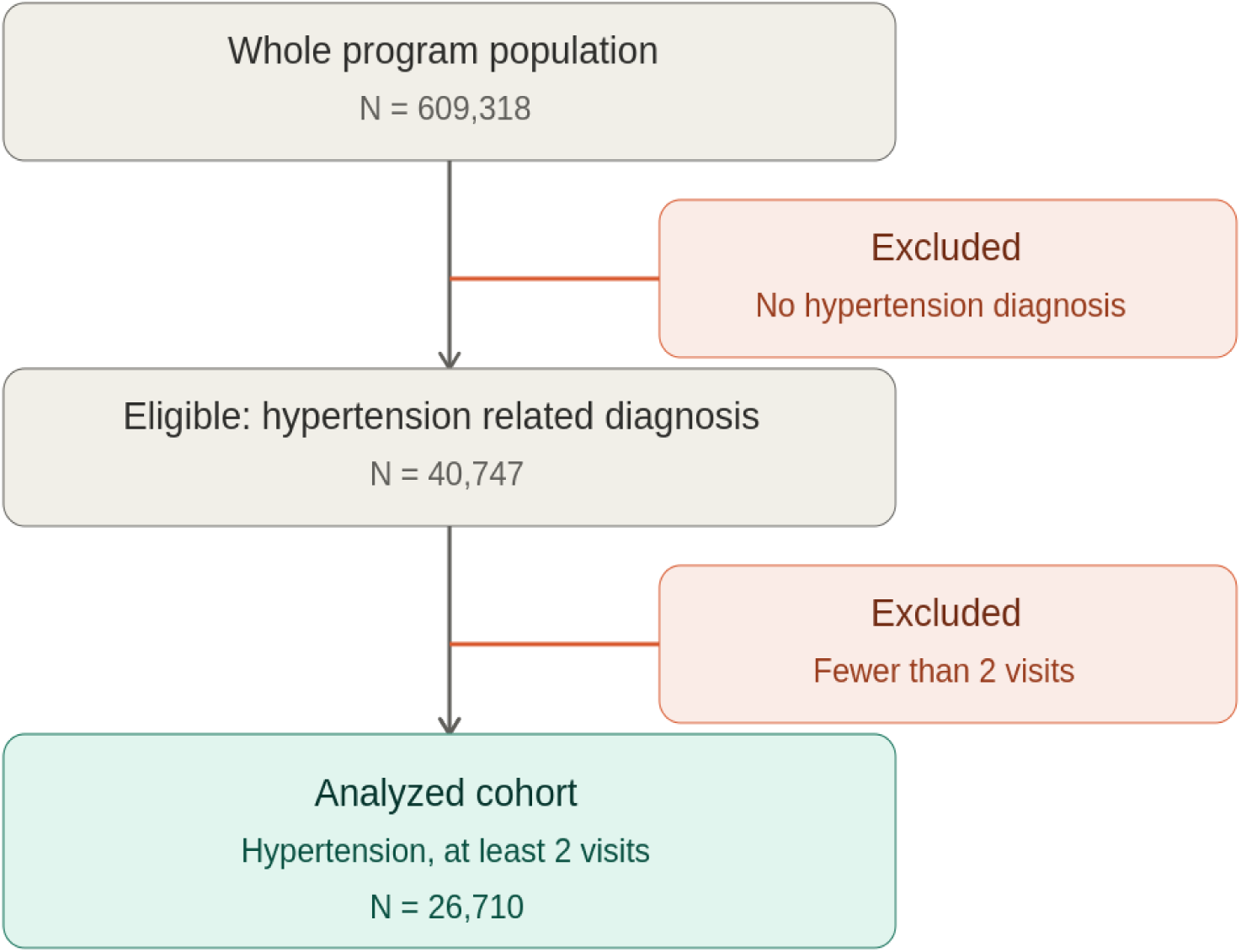
Cohort selection flow.

### Visit-Based Retention Patterns

Observed EHR contact declined progressively over follow-up. The proportion of patients with at least one recorded encounter decreased from 70.4% at 6 months to 51.8% at 12 months, 22.1% at 24 months, and 8.9% at 36 months. Patients had a median of 4 visits (IQR, 2–7), a median follow-up of 377 days (IQR, 139–685), and a median inter-visit interval of 35 days (IQR, 1–105).

Among patients who experienced prolonged interruptions in care, the median time to the first inter-visit gap was 276 days (IQR, 215–382) for gaps >6 months, 468 days (IQR, 402–604) for gaps >12 months, and 684 days (IQR, 607–799) for gaps >18 months. Patients with only two visits or follow-up shorter than the specified gap threshold were excluded from these estimates.

BP control rates among patients with available measurements remained relatively stable over time. At 12 months, 16,101 patients remained under observation, of whom 11,692 (72.6%) had a BP measurement and 7,053 (60.3%) achieved BP control, representing 26.4% of the full cohort. Corresponding values at 24 months were 7,513, 5,565 (74.1%), and 3,337 (60.0%) (12.5% of the cohort), respectively; at 36 months, they were 3,100, 2,322 (74.9%), and 1,373 (59.1%) (5.1% of the cohort) **(Table 1).**

**Table 1.**
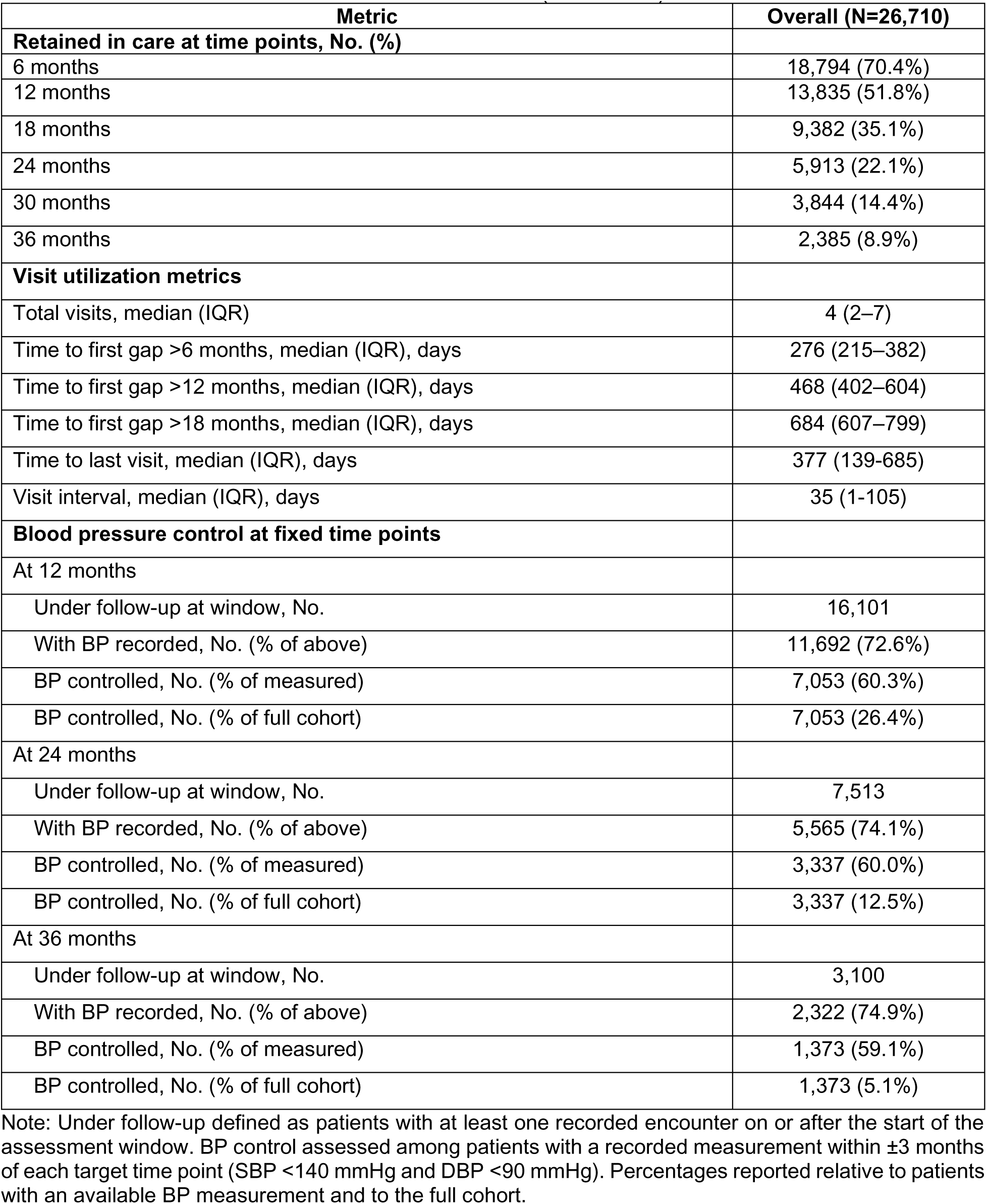
Overall Visit-Based Retention Patterns (N=26,710)

### Latent class trajectory model (1-month interval)

#### Model selection

The 1-month model was specified as the primary analysis, with the 3-month model used to assess robustness to temporal resolution. Models with one to six latent classes were evaluated using the Bayesian Information Criterion (BIC), GRoLTS quality metrics, and clinical interpretability (**Supplementary Table S1**). Although BIC decreased monotonically, the improvement diminished beyond four classes. The four-class solution satisfied all GRoLTS criteria (relative entropy = 0.821, minimum APP = 0.770, minimum OCC = 6.49), whereas the five-class model showed poorer class separation (relative entropy = 0.714; minimum OCC = 4.84). Accordingly, the four-class model was selected based on model fit, classification quality, and interpretability.

Bootstrap validation (200 resamples) demonstrated excellent classification stability (100% convergence). The Adjusted Rand Index (ARI) had a mean of 0.969 (median 0.972, SD 0.008; 95% percentile interval 0.953–0.978), indicating highly reproducible class assignments.

Four distinct patterns of observed EHR contact were identified (**Figure 2**). The Low Observed Follow-up group (n = 18,308; 68.5%) showed a rapid decline in contact after the index visit, approaching zero by month 27. The Stable Low Observed Retention group (n = 4,879; 18.3%) declined sharply after the first month but subsequently maintained a stable attendance probability of approximately 0.2 throughout follow-up. The Delayed Decline in Observed Visits group (n = 2,945; 11.0%) maintained moderate contact for approximately 12 months before declining gradually to near zero by month 27. Around month 19, its trajectory crossed that of the Stable Low Observed Retention group, which thereafter maintained the higher level of observed contact. The Sustained Observed Retention group (n = 578; 2.2%) maintained the highest level of contact throughout follow-up, remaining at approximately 0.3 by month 36.

**Figure 2.**
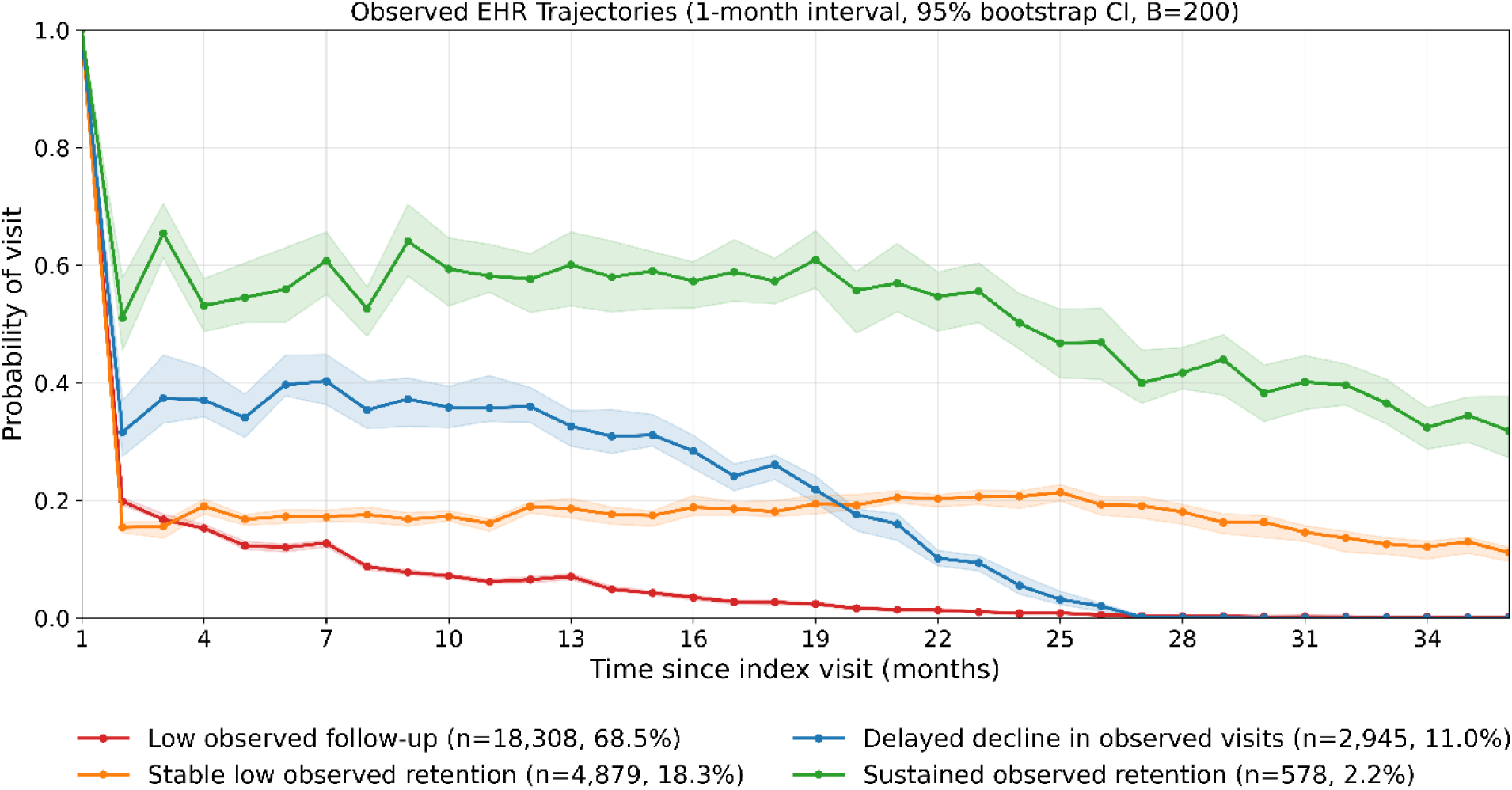
Observed EHR contact trajectories by 1-month interval. Trajectories represent the probability of having at least one visit within each 1-month interval following the index visit. The first interval (0 to 1 months, labeled as 1 month) includes the index visit and therefore reflects initial attendance.

#### Baseline characteristics

Baseline characteristics differed significantly across trajectory groups (all p < 0.001; **Table 2**). The Sustained Observed Retention group was the oldest (mean age 73.4 years), followed by the Delayed Decline in Observed Visits group (71.5 years), whereas the Low Observed Follow-up and Stable Low Observed Retention groups had similar mean ages (68.4 and 68.6 years). The Stable Low Observed Retention group had the lowest proportion of women (15.4%), while the remaining groups ranged from 22.5% to 25.5%.

**Table 2.**
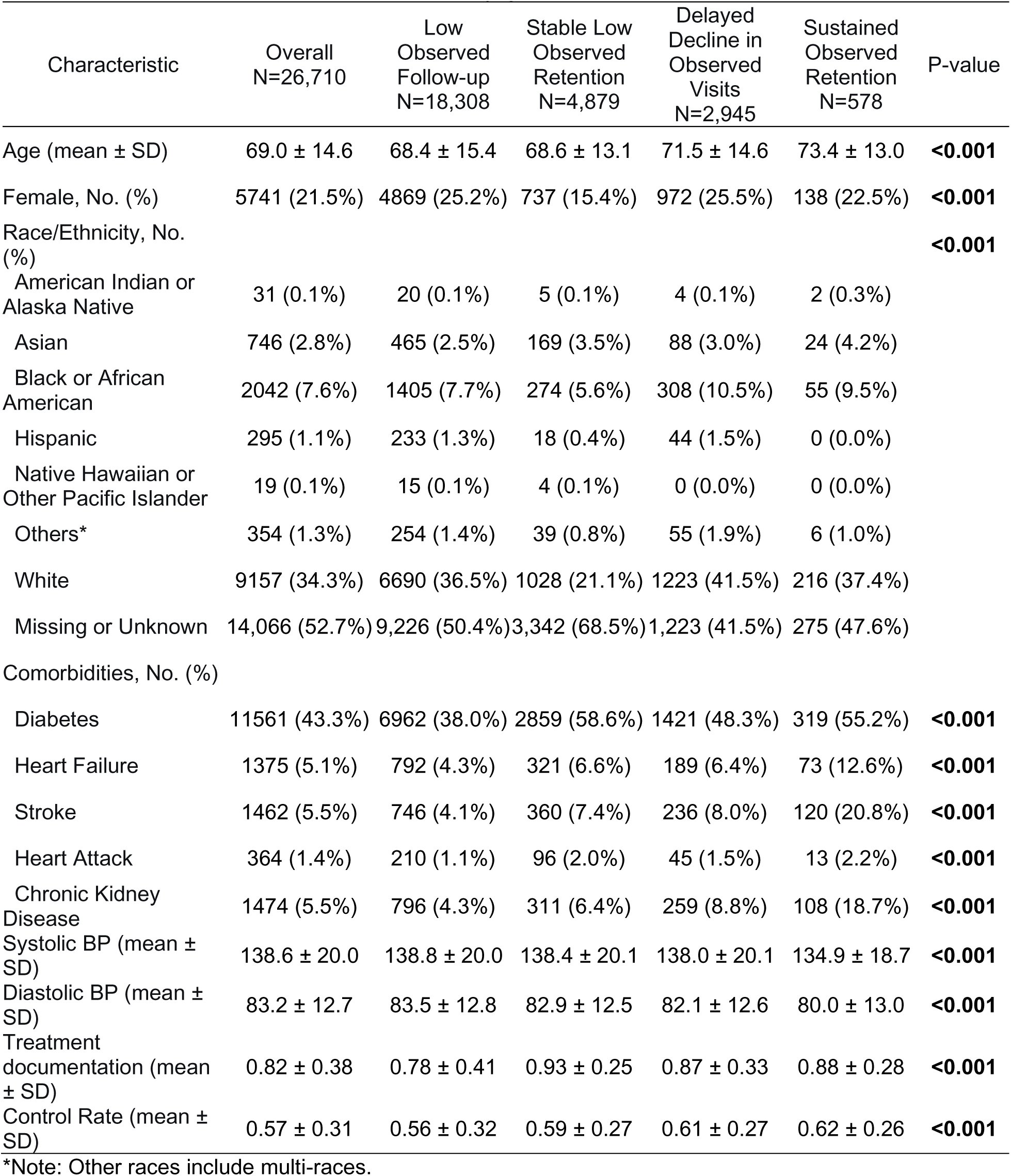
1-month interval characteristics by group.

Comorbidity burden varied substantially across groups. The Sustained Observed Retention group had the highest prevalence of heart failure (12.6%), stroke (20.8%), and chronic kidney disease (18.7%), whereas diabetes was most prevalent in the Stable Low Observed Retention group (58.6%) and least common in the Low Observed Follow-up group (38.0%). Baseline BP differed statistically but only minimally between groups. Standardized mean differences (SMDs) for systolic BP ranged from −0.023 to −0.205 and for diastolic BP from −0.046 to −0.274, indicating negligible to small differences relative to the Low Observed Follow-up group.

Treatment-related measures showed greater between-group variation. The Stable Low Observed Retention group had the highest treatment documentation rate (0.93 ± 0.25), followed by the Sustained Observed Retention (0.88 ± 0.28) and Delayed Decline in Observed Visits (0.87 ± 0.33) groups, while the Low Observed Follow-up group had the lowest rate (0.78 ± 0.41). BP control was highest in the Sustained Observed Retention group (0.62 ± 0.26), followed by the Delayed Decline in Observed Visits (0.61 ± 0.27) and Stable Low Observed Retention (0.59 ± 0.27) groups, and lowest in the Low Observed Follow-up group (0.56 ± 0.32).

### Latent class trajectory model (3-month interval)

#### Model selection

The 3-month model identified patterns of observed EHR contact that were broadly consistent with those from the primary 1-month analysis. Models with two to six latent classes were evaluated using the Bayesian Information Criterion (BIC), GRoLTS quality metrics, and clinical interpretability (**Supplementary Table S2**). Although BIC decreased with increasing numbers of classes, the improvement diminished beyond four classes. The four-class solution satisfied all GRoLTS criteria (relative entropy = 0.771, minimum APP = 0.778, minimum OCC = 5.39), whereas the five-class model showed poorer class separation (relative entropy = 0.700; minimum OCC = 4.66). Accordingly, the four-class model was selected.

Bootstrap validation (200 resamples) demonstrated excellent classification stability (100% convergence), with a mean ARI of 0.966 (median 0.972, SD 0.018; 95% percentile interval 0.930–0.991). Four trajectory groups were identified (**Figure 3**). The Low Observed Follow-up group (n = 18,008; 67.4%) showed a rapid, sustained decline in observed contact after the index visit. The Delayed Decline in Observed Visits group (n = 3,892; 14.6%) maintained high contact through approximately 18 months before declining sharply. The Stable Low Observed Retention group (n = 3,320; 12.4%) declined initially but subsequently increased to a moderate, sustained level of contact, overtaking the Delayed Decline in Observed Visits group between months 21 and 24. The Sustained Observed Retention group (n = 1,490; 5.6%) maintained the highest probability of observed contact throughout follow-up, remaining approximately 0.6 at 36 months.

**Figure 3.**
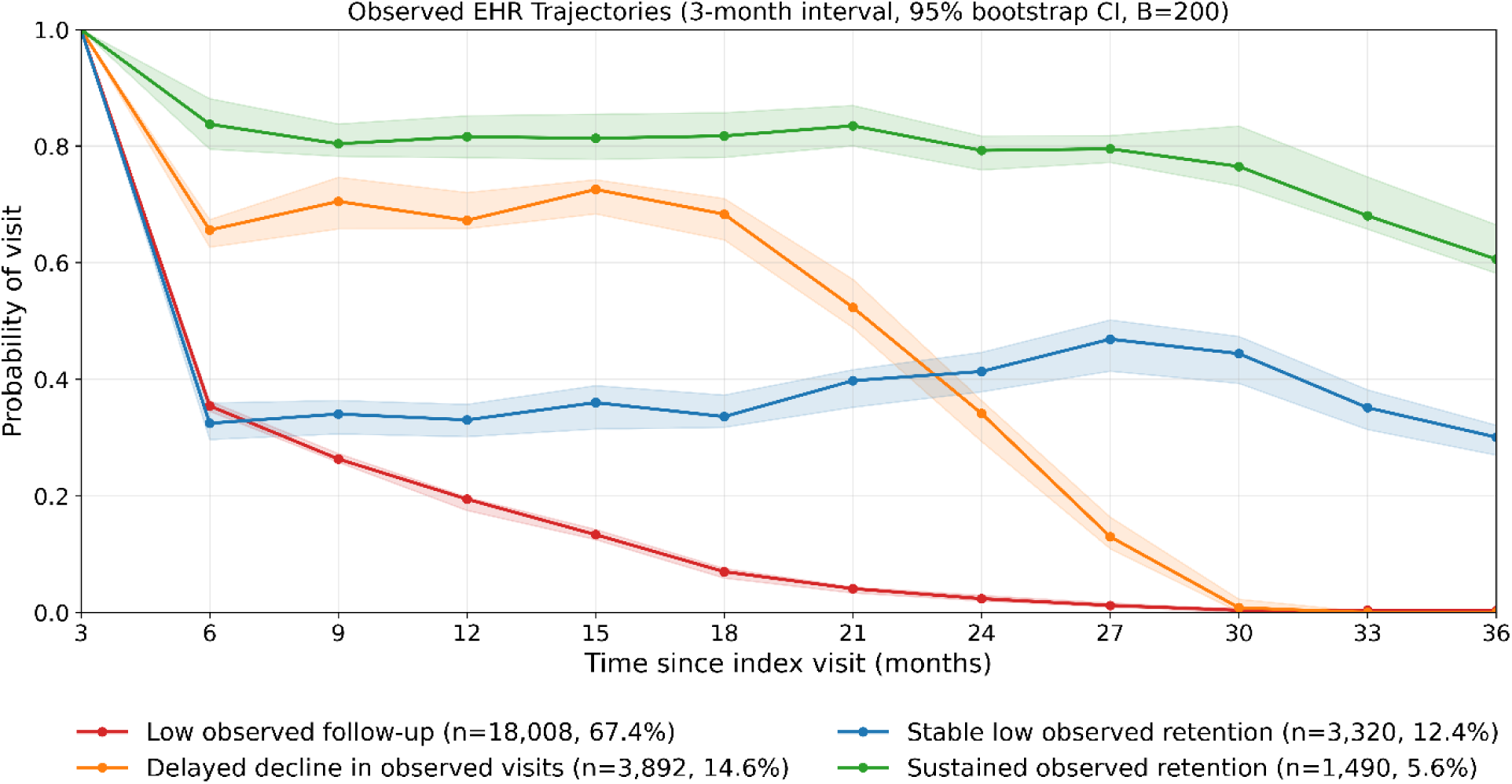
Observed EHR contact trajectories by 3-month interval. Trajectories represent the probability of having at least one visit within each 3-month interval following the index visit. The first interval (0 to 3 months, labeled as 3 month) includes the index visit and therefore reflects initial attendance.

#### Baseline characteristics

Baseline characteristics were broadly consistent with those observed in the 1-month model, with significant differences across trajectory groups (all p < 0.001; **Table 3**). The Delayed Decline in Observed Visits group had the highest mean age (71.7 years), whereas the Stable Low Observed Retention group was the youngest (67.8 years). The proportion of women was highest in the Low Observed Follow-up group (22.6%) and lowest in the Sustained Observed Retention group (13.6%).

**Table 3.**
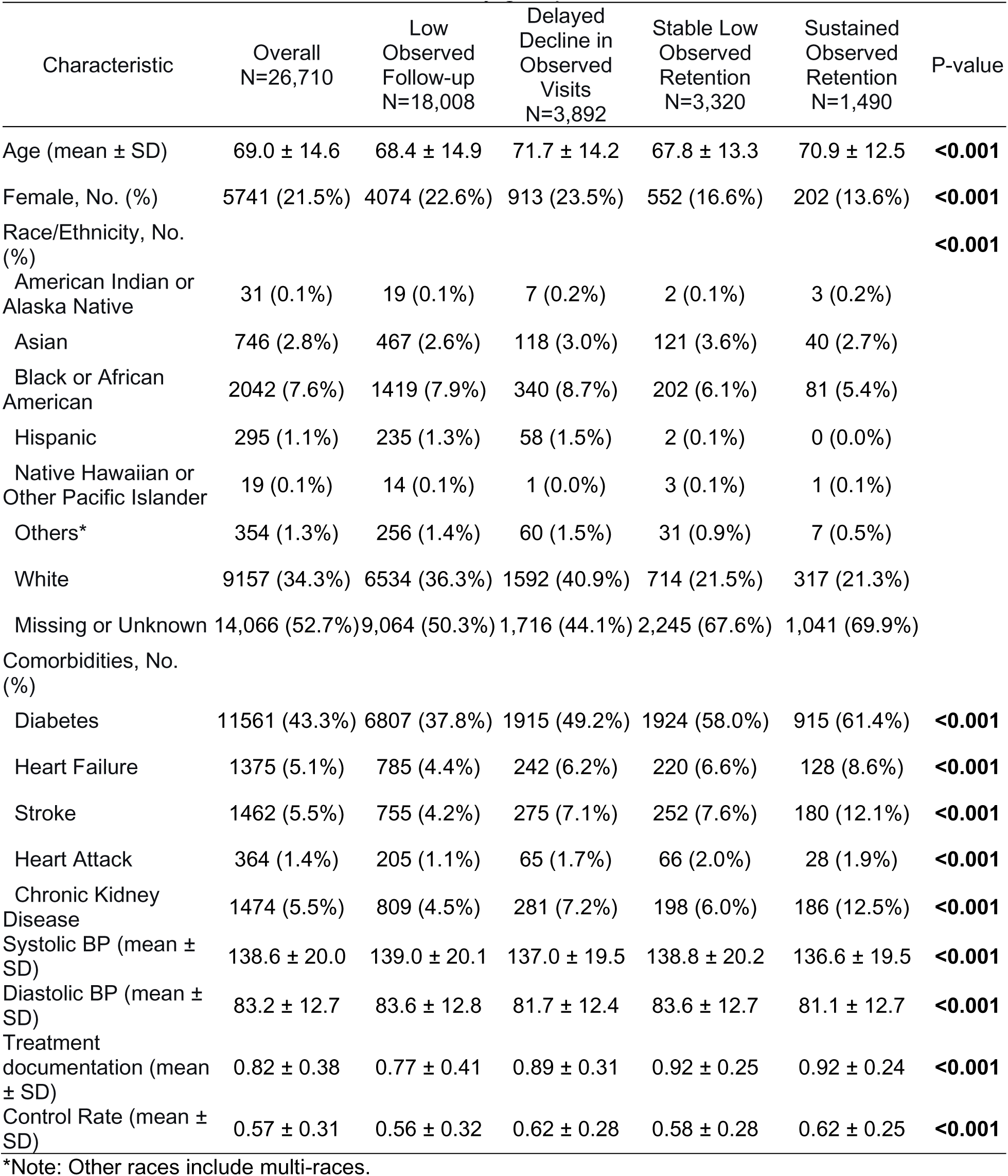
3-month interval characteristics by group.

| Characteristic | Overall<br>N=26,710 | Low<br>Observed<br>Follow-up<br>N=18,008 | Delayed<br>Decline in<br>Observed<br>Visits<br>N=3,892 | Stable Low<br>Observed<br>Retention<br>N=3,320 | Sustained<br>Observed<br>Retention<br>N=1,490 | P-value |
| --- | --- | --- | --- | --- | --- | --- |
| Age (mean ± SD) | 69.0 ± 14.6 | 68.4 ± 14.9 | 71.7 ± 14.2 | 67.8 ± 13.3 | 70.9 ± 12.5 | **<0.001** |
| Female, No. (%) | 5741 (21.5%) | 4074 (22.6%) | 913 (23.5%) | 552 (16.6%) | 202 (13.6%) | **<0.001** |
| Race/Ethnicity, No. (%) |  |  |  |  |  | **<0.001** |
| American Indian or Alaska Native | 31 (0.1%) | 19 (0.1%) | 7 (0.2%) | 2 (0.1%) | 3 (0.2%) |  |
| Asian | 746 (2.8%) | 467 (2.6%) | 118 (3.0%) | 121 (3.6%) | 40 (2.7%) |  |
| Black or African American | 2042 (7.6%) | 1419 (7.9%) | 340 (8.7%) | 202 (6.1%) | 81 (5.4%) |  |
| Hispanic | 295 (1.1%) | 235 (1.3%) | 58 (1.5%) | 2 (0.1%) | 0 (0.0%) |  |
| Native Hawaiian or Other Pacific Islander | 19 (0.1%) | 14 (0.1%) | 1 (0.0%) | 3 (0.1%) | 1 (0.1%) |  |
| Others* | 354 (1.3%) | 256 (1.4%) | 60 (1.5%) | 31 (0.9%) | 7 (0.5%) |  |
| White | 9157 (34.3%) | 6534 (36.3%) | 1592 (40.9%) | 714 (21.5%) | 317 (21.3%) |  |
| Missing or Unknown | 14,066 (52.7%) | 9,064 (50.3%) | 1,716 (44.1%) | 2,245 (67.6%) | 1,041 (69.9%) |  |
| Comorbidities, No. (%) |  |  |  |  |  |  |
| Diabetes | 11561 (43.3%) | 6807 (37.8%) | 1915 (49.2%) | 1924 (58.0%) | 915 (61.4%) | **<0.001** |
| Heart Failure | 1375 (5.1%) | 785 (4.4%) | 242 (6.2%) | 220 (6.6%) | 128 (8.6%) | **<0.001** |
| Stroke | 1462 (5.5%) | 755 (4.2%) | 275 (7.1%) | 252 (7.6%) | 180 (12.1%) | **<0.001** |
| Heart Attack | 364 (1.4%) | 205 (1.1%) | 65 (1.7%) | 66 (2.0%) | 28 (1.9%) | **<0.001** |
| Chronic Kidney Disease | 1474 (5.5%) | 809 (4.5%) | 281 (7.2%) | 198 (6.0%) | 186 (12.5%) | **<0.001** |
| Systolic BP (mean ± SD) | 138.6 ± 20.0 | 139.0 ± 20.1 | 137.0 ± 19.5 | 138.8 ± 20.2 | 136.6 ± 19.5 | **<0.001** |
| Diastolic BP (mean ± SD) | 83.2 ± 12.7 | 83.6 ± 12.8 | 81.7 ± 12.4 | 83.6 ± 12.7 | 81.1 ± 12.7 | **<0.001** |
| Treatment documentation (mean ± SD) | 0.82 ± 0.38 | 0.77 ± 0.41 | 0.89 ± 0.31 | 0.92 ± 0.25 | 0.92 ± 0.24 | **<0.001** |
| Control Rate (mean ± SD) | 0.57 ± 0.31 | 0.56 ± 0.32 | 0.62 ± 0.28 | 0.58 ± 0.28 | 0.62 ± 0.25 | **<0.001** |

\*Note: Other races include multi-races.

The Sustained Observed Retention group had the greatest comorbidity burden, including the highest prevalence of diabetes (61.4%), stroke (12.1%), and chronic kidney disease (12.5%). Heart failure prevalence increased progressively across the Low Observed Follow-up, Delayed Decline in Observed Visits, Stable Low Observed Retention, and Sustained Observed Retention groups (4.4%, 6.2%, 6.6%, and 8.6%, respectively). Baseline BP differed minimally between groups. Standardized mean differences for systolic BP ranged from −0.011 to −0.123 and for diastolic BP from 0.005 to −0.196, indicating negligible differences relative to the Low Observed Follow-up group. Treatment-related measures showed greater variation. Treatment documentation rates were highest in the Stable Low Observed Retention and Sustained Observed Retention groups (both 0.92), followed by the Delayed Decline in Observed Visits group (0.89), and lowest in the Low Observed Follow-up group (0.77). BP control rates were highest in the Delayed Decline in Observed Visits and Sustained Observed Retention groups (both 0.62), intermediate in the Stable Low Observed Retention group (0.58), and lowest in the Low Observed Follow-up group (0.56).

### First-to-last visit comparisons

Across both trajectory models, systolic BP (SBP) and diastolic BP (DBP) were lower at the last than the first recorded visit (**Figure 4**). In the 1-month model, mean SBP reductions ranged from 4.7 to 5.4 mmHg across groups and were statistically significant except in the Sustained Observed Retention group (2.3 mmHg; p = 0.068). DBP declined significantly in all groups, with reductions of 3.3–4.2 mmHg. In the 3-month model, both SBP (3.8–5.5 mmHg) and DBP (3.4–4.0 mmHg) decreased significantly across all groups (all p < 0.001).

**Figure 4a.**
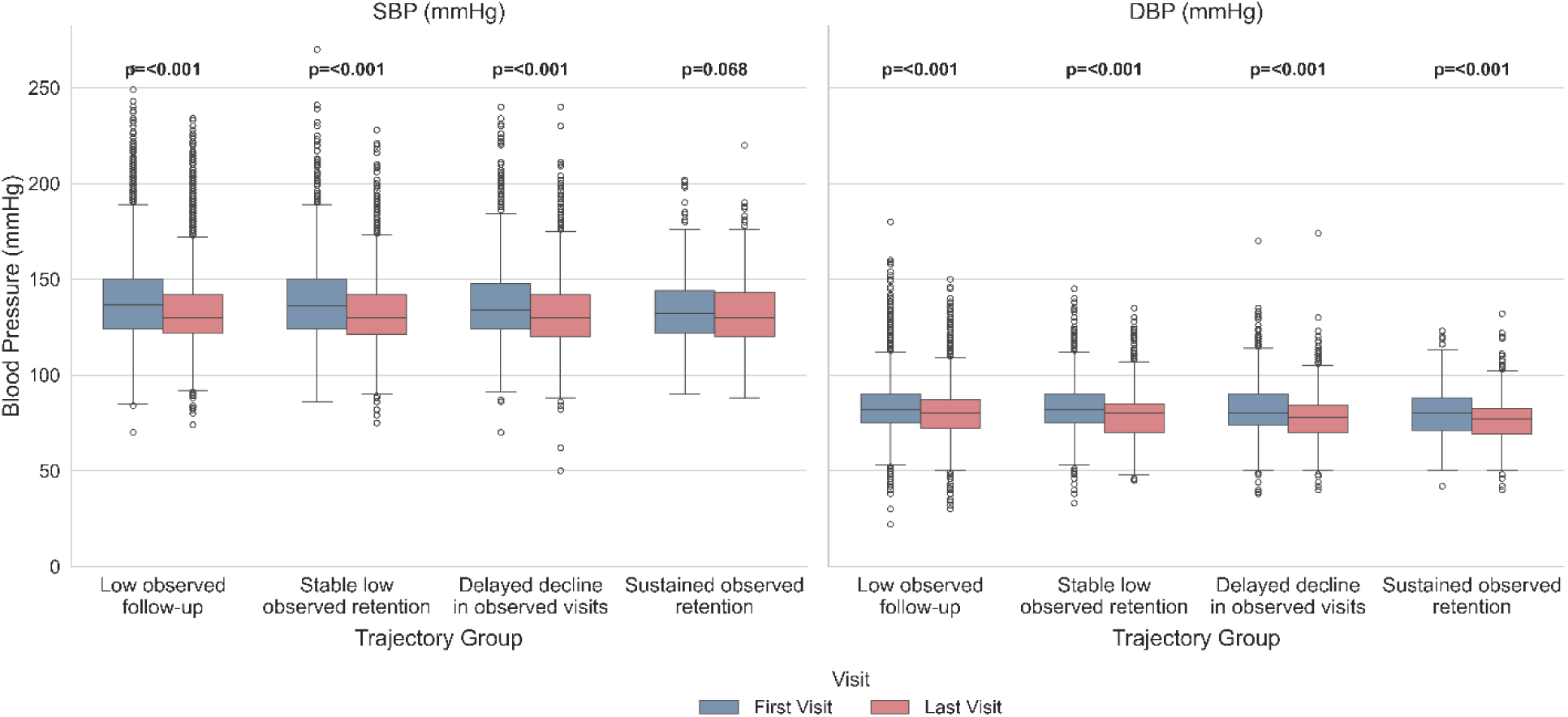
Blood pressure at first and last visit across trajectory groups (1-month)

**Figure 4b.**
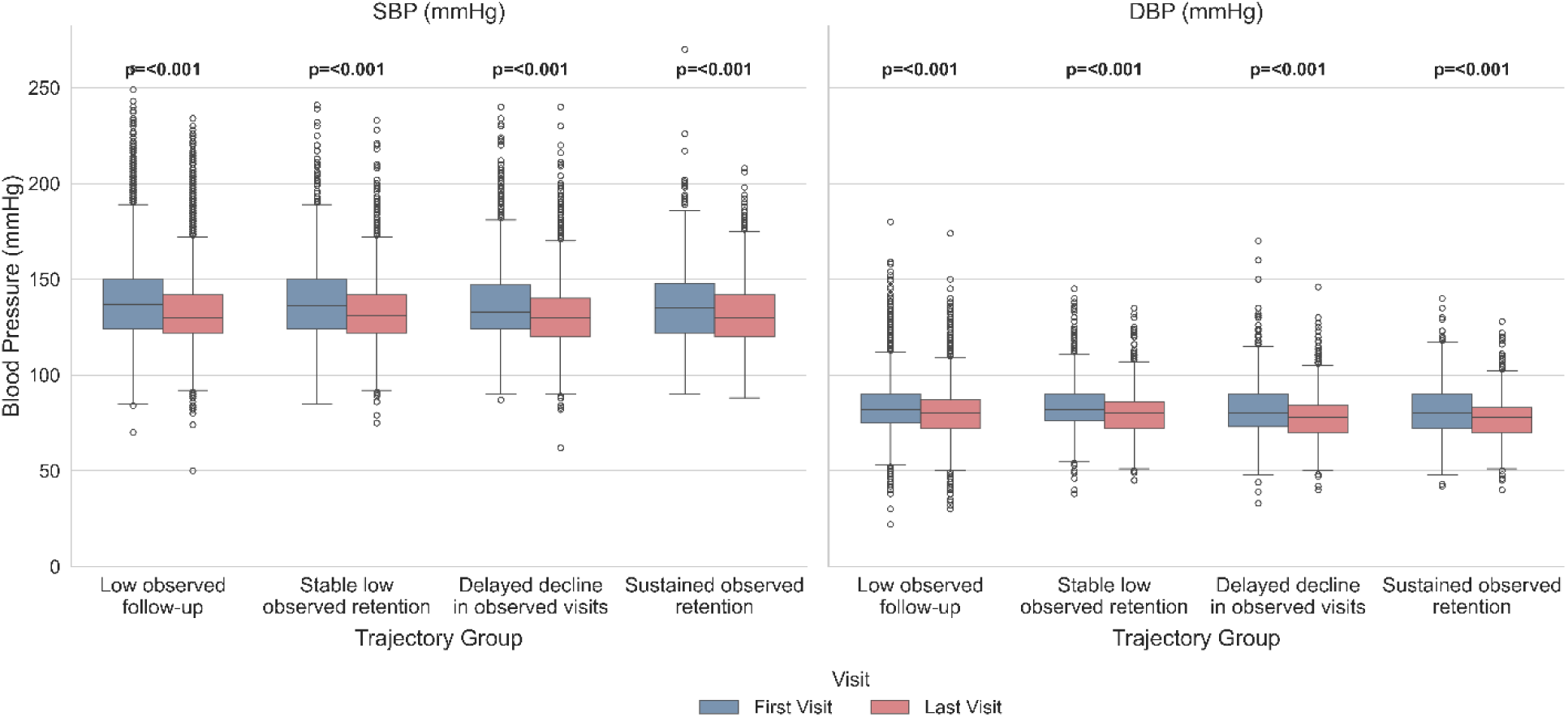
Blood pressure at first and last visit across trajectory groups (3-month)

The smaller SBP reduction in the Sustained Observed Retention group should be interpreted cautiously. Because this group had the longest observed follow-up, first-to-last comparisons span a longer observation window and are influenced by differential follow-up duration rather than reflecting a greater rate of BP improvement. Treatment documentation and BP control increased from the first to the last visit in both models (Figure 5). In the 1-month model, treatment documentation increased least in the Low Observed Follow-up group (77.3% to 78.3%) and most in the Sustained Observed Retention group (79.1% to 95.0%) (all p < 0.001). Similar findings were observed in the 3-month model. BP control also improved significantly across nearly all groups; the only exception was the Sustained Observed Retention group in the 1-month model, where the increase (58.1% to 63.6%) did not reach statistical significance (p = 0.085). In the 3-month model, BP control increased significantly in all groups (all p < 0.001).

**Figure 5a.**
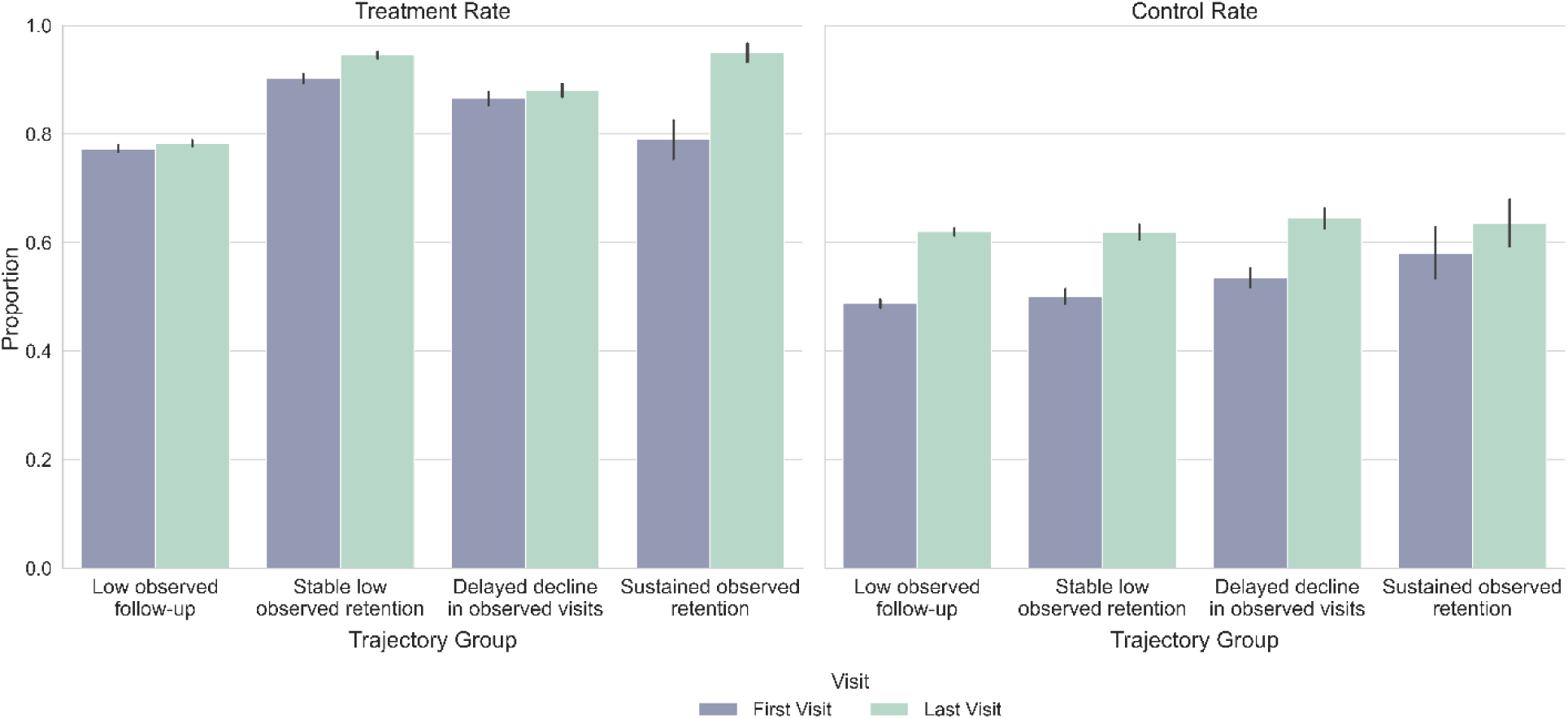
Treatment and control rates at first vs last visit by trajectory group (1-month)

**Figure 5b.**
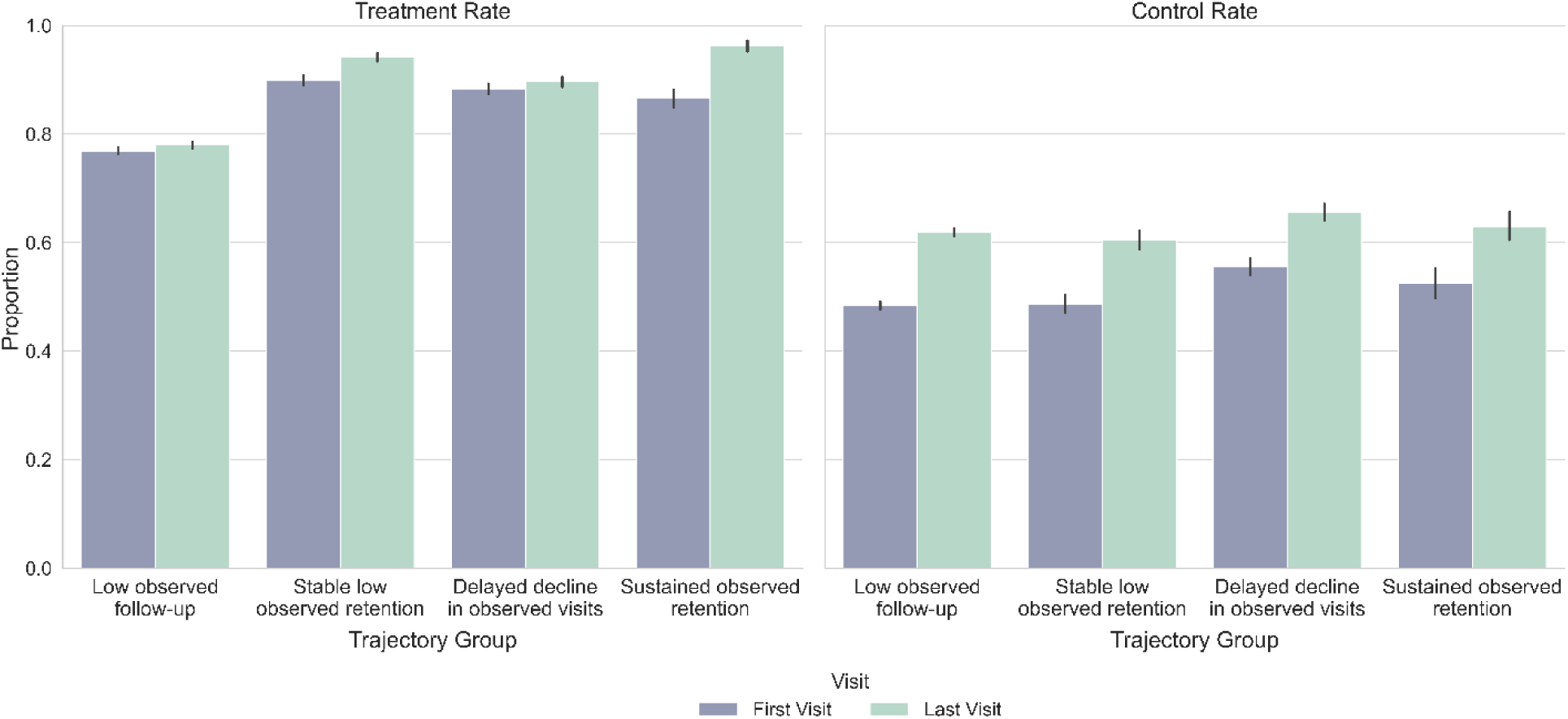
Treatment and control rates at first vs last visit by trajectory group (3-month)

### Fixed time-point BP outcomes

To reduce bias arising from differing observation windows, BP was also evaluated at fixed intervals (12, 24, and 36 months) using the nearest measurement within a ±3-month window (**Supplementary Figures S1–S2; Tables S4–S5**). Because analyses were restricted to patients with available BP measurements, results are conditional on continued observed contact and should not be interpreted as population-level estimates. In the 1-month model, SBP differed significantly across trajectory groups at 12, 24, and 36 months (p < 0.05), although absolute differences were modest (≤2 mmHg at most time points). The Low Observed Follow-up group generally had the lowest SBP, whereas the Stable Low Observed Retention and Sustained Observed Retention groups tended to have the highest. DBP differed little between groups.

In the 3-month model, SBP differed significantly at 12 and 24 months but not at 36 months. The Stable Low Observed Retention group consistently had the highest SBP at 12 and 24 months, while values converged across all groups by 36 months. DBP differences remained small throughout follow-up. Importantly, the Sustained Observed Retention group did not exhibit the lowest SBP at any fixed time point despite appearing to have greater BP improvement in first-to-last comparisons. Because fixed time-point analyses compare patients at equivalent follow-up durations, they provide a more appropriate assessment of BP differences across trajectory groups. Overall, these findings suggest that trajectory group membership is more strongly associated with care process measures, including treatment documentation and BP control, than with absolute BP levels.

### Adjusted models: treatment documentation rate and control rate

After adjustment for age, sex, race/ethnicity, baseline systolic BP, diabetes, heart failure, stroke, and chronic kidney disease, all non-reference trajectory groups had significantly higher treatment documentation rates than the Low Observed Follow-up group in both temporal resolutions. In the 1-month model, the largest adjusted differences were observed in the Sustained Observed Retention (β = 0.176, 95% CI 0.141–0.211), Stable Low Observed Retention (β = 0.164, 95% CI 0.153–0.176), and Delayed Decline in Observed Visits (β = 0.080, 95% CI 0.066–0.094) groups (all p < 0.001). Similar results were obtained in the 3-month model, with the largest effects in the Sustained Observed Retention (β = 0.192, 95% CI 0.171–0.212), Delayed Decline in Observed Visits (β = 0.163, 95% CI 0.150–0.177), and Stable Low Observed Retention (β = 0.099, 95% CI 0.086–0.111) groups (all p < 0.001). These findings indicate that the association between trajectory group and treatment documentation was robust to adjustment for baseline characteristics.

Adjusted associations with BP control were smaller and less consistent. In the 1-month model, BP control was higher in the Delayed Decline in Observed Visits (β = 0.030, 95% CI 0.020–0.040) and Stable Low Observed Retention (β = 0.015, 95% CI 0.007–0.023) groups (both p < 0.001), whereas the association for the Sustained Observed Retention group was not significant (β = 0.010, 95% CI −0.014 to 0.035; p = 0.417). In the 3-month model, the largest adjusted difference was observed in the Stable Low Observed Retention group (β = 0.035, 95% CI 0.026–0.044), followed by the Sustained Observed Retention group (β = 0.030, 95% CI 0.016–0.045) (both p < 0.001), while the Delayed Decline in Observed Visits group showed only borderline significance (β = 0.010, 95% CI 0.000–0.019; p = 0.052). Overall, adjustment had little effect on the association between trajectory group and treatment documentation but attenuated associations with BP control, particularly for the Sustained Observed Retention group. These findings suggest that differences in BP control are partly explained by baseline case mix and comorbidity burden rather than observed contact alone.

### Concordance between 1-month and 3-month models

Individual-level trajectory assignments showed moderate agreement between the 1- month and 3-month models (Adjusted Rand Index = 0.765; Cohen’s κ = 0.389). The lower κ likely reflects the marked imbalance in class sizes, with the Low Observed Follow-up group comprising approximately two-thirds of the cohort. Concordance varied by phenotype. The Low Observed Follow-up (94.8%) and Sustained Observed Retention (85.1%) groups showed high agreement across temporal resolutions, followed by the Delayed Decline in Observed Visits group (80.6%). The Stable Low Observed Retention group showed the lowest concordance (62.1%), with many patients reassigned to the Sustained Observed Retention (20.1%) or Delayed Decline in Observed Visits (15.5%) groups in the 3-month model, suggesting greater sensitivity to temporal aggregation. Despite these differences, both temporal resolutions consistently identified the same four trajectory phenotypes. The high concordance of the Low Observed Follow-up and Sustained Observed Retention groups supports the robustness of the latent class solution, whereas the intermediate phenotypes appear more dependent on the choice of time interval.

### Sensitivity Analysis

Given substantial right censoring, a sensitivity analysis restricted the cohort to patients with at least 24 months of observed follow-up (n = 5,567; 20.8% of the full cohort). Censoring varied markedly across trajectory groups. In the 1-month model, censoring was highest in the Low Observed Follow-up (97.2%; median last observation, 5.7 months) and Delayed Decline in Observed Visits (95.9%; median, 18.1 months) groups, compared with the Stable Low Observed Retention (9.7%; median, 30.6 months) and Sustained Observed Retention (13.1%; median, 34.0 months) groups. Similar patterns were observed in the 3-month model, with substantial censoring in the Low Observed Follow-up (98.0%; median, 5.6 months) and Stable Low Observed Retention (88.0%; median, 19.6 months) groups, but minimal censoring in the Delayed Decline in Observed Visits (3.9%; median, 30.7 months) and Sustained Observed Retention (0.0%; median, 33.6 months) groups. These findings indicate that lower observed contact in some trajectory groups may partly reflect shorter administrative observation windows rather than confirmed loss to follow-up.

Within the restricted cohort, a four-class solution remained the most supported model in both temporal resolutions, although class separation was reduced compared with the primary analysis. In the 1-month sensitivity model, BIC showed a relative improvement at K = 4 (reduction of 265.2 from K = 3), with diminishing gains for additional classes. The K = 4 model demonstrated moderate classification quality (relative entropy = 0.737, minimum APP = 0.753, minimum OCC = 4.49). In the 3-month sensitivity model, K = 4 similarly represented the point of diminishing improvement (BIC = 79,240.5; reduction of 277.3 from K = 3), with acceptable APP and OCC values (minimum APP = 0.741; minimum OCC = 7.23), although entropy remained below the recommended threshold (0.648). Thus, K = 4 was retained based on the BIC pattern, consistency with the primary analysis, and clinical interpretability rather than complete GRoLTS threshold compliance. The sensitivity analysis supports the stability of the four-group structure among patients with longer observable follow-up, while highlighting reduced model separation due to the smaller sample size and the influence of administrative censoring on observed contact trajectories.

## DISCUSSION

Latent class trajectory modelling of repeated binary visit indicators consistently identified four observed EHR contact phenotypes across both temporal resolutions, with high bootstrap assignment stability (200 resamples). The four-class structure was also supported in a sensitivity analysis restricted to patients with ≥24 months of observed follow-up. Cross-resolution comparison demonstrated moderate agreement between individual-level assignments (ARI = 0.765; Cohen’s κ = 0.389). These findings support internal reproducibility but do not establish the optimal number of classes, exclude censoring-related artefacts, or provide external validation. Because bootstrap resampling preserves the underlying observation process, it cannot address bias introduced by variable administrative follow-up windows. The most distinct phenotypes were the most robust across temporal resolutions. The Low Observed Follow-up group (67–68% of the cohort) and Sustained Observed Retention group (2–6%) showed high concordance between models (94.8% and 85.1%, respectively). In contrast, the intermediate phenotypes were more sensitive to interval specification. The Delayed Decline in Observed Visits and Stable Low Observed Retention groups showed lower concordance (80.6% and 62.1%, respectively), with reassignment of a proportion of Stable Low Observed Retention patients to higher-retention categories in the 3-month model. This likely reflects smoothing of short-term visit variability by longer aggregation intervals and warrants caution when interpreting these intermediate trajectories.

A key finding was the crossover between the Delayed Decline in Observed Visits and Stable Low Observed Retention phenotypes. In the 1-month model, the Delayed Decline group initially demonstrated higher observed contact but declined below the Stable Low Observed Retention group at approximately month 19. A similar pattern was observed in the 3-month model, with crossover occurring between months 21 and 24 after a more pronounced initial decline. Despite differences in timing, both models demonstrate that early observed contact does not reliably predict long-term retention patterns. Patients with initially high observed contact may subsequently experience substantial decline. This finding aligns with evidence that chronic disease care utilization and adherence-related behaviors are dynamic over time.[15] Prior trajectory studies in other chronic conditions have shown that non-monotonic patterns of adherence and care engagement are associated with clinically relevant outcomes, including cardiovascular risk.[16] Studies of statin and antihypertensive medication adherence similarly demonstrate that declines from initially high adherence patterns are associated with adverse outcomes, even among patients with comparable baseline status.[17, 18] Extending this concept to visit-based observed contact, our findings highlight the limitation of using early follow-up encounters as a surrogate for long-term retention and support longitudinal monitoring of contact patterns rather than assessment at a single time point.

The primary limitation in interpreting trajectory shapes is substantial administrative right censoring. Overall, 79.2% of patients had <24 months of observed follow-up, with censoring concentrated among the lowest-contact phenotypes. In the 1-month model, 97.2% of the Low Observed Follow-up group and 95.9% of the Delayed Decline in Observed Visits group were censored, compared with 9.7% and 13.1% in the Stable Low Observed Retention and Sustained Observed Retention groups, respectively. Similar patterns were observed in the 3-month model. Therefore, declines toward near-zero observed contact in highly censored groups likely reflect administrative observation limits rather than confirmed disengagement from care. The sensitivity analysis restricted to patients with ≥24 months of follow-up provides only partial reassurance because selection into this subgroup is itself related to observed care patterns. Although a four-class structure remained plausible, weaker separation metrics indicate reduced stability when observation windows are more comparable. Moreover, this subgroup represents a selected minority of the cohort. Future studies should distinguish administrative censoring from true reductions in care through approaches such as complete observation-window designs, inverse-probability-of-censoring weighting, joint modelling of visit and censoring processes, and external validation in independent populations before applying these phenotypes for intervention targeting.[19]

Clinical associations did not support a simple relationship between higher observed contact and better BP outcomes. Baseline BP did not meaningfully differentiate trajectory groups. Although between-group differences in SBP and DBP were statistically significant, all standardized mean differences were <0.2, indicating negligible clinical differences. The Sustained Observed Retention group had the lowest baseline BP despite the highest comorbidity burden, suggesting that higher observed contact was more closely related to management complexity and comorbid conditions, including diabetes, heart failure, stroke, and chronic kidney disease, than to baseline hypertension severity.

Treatment documentation demonstrated a more consistent association with observed contact trajectories than BP control. Unadjusted analyses showed higher treatment documentation and control rates among higher-contact groups. After adjustment for demographic factors, baseline BP, and comorbidities, treatment documentation remained significantly higher across all non-reference groups in both temporal models, supporting a robust association between observed contact trajectory and recorded treatment coverage. In contrast, adjusted differences in BP control were smaller and less consistent. The Sustained Observed Retention group showed no significant adjusted difference in the 1-month model (β = 0.010, 95% CI −0.014 to 0.035; p = 0.417), suggesting that its higher unadjusted control rate was partly attributable to differences in baseline characteristics and comorbidity burden. These findings are consistent with confounding by indication: patients with greater clinical complexity may have both greater observed contact and more intensive management.

Fixed time-point BP analyses further demonstrate the limitations of first-to-last comparisons. Although the Sustained Observed Retention group showed the largest apparent SBP reduction in paired analyses, it did not have the lowest SBP at any fixed follow-up time point. By 36 months, between-group SBP differences were no longer significant in the 3-month model, and group rankings varied across time points. Because the Sustained Observed Retention group had substantially longer observed follow-up, first-to-last comparisons capture differences in observation duration rather than treatment response alone. Given its low baseline BP, regression to the mean is unlikely to explain this pattern. Fixed time-point analyses therefore provide a more appropriate comparison and suggest that observed contact trajectories are more strongly associated with care process measures than with absolute BP levels.

These findings are consistent with intensive BP management trials, including SPRINT trial and STEP trial, which demonstrated cardiovascular benefits from sustained monitoring and treatment intensification despite relatively modest differences in achieved BP levels[20, 21]. Whether the observed association between contact trajectories and treatment documentation translates into differences in cardiovascular outcomes requires future linkage to clinical endpoints, analogous to prior work evaluating medication adherence trajectories[6, 18].

### Limitations

There are several limitations in this study. First, healthcare utilization outside the observed system was not captured, potentially underestimating true visit frequency. Second, binary visit indicators measure observed contact only and do not capture visit quality, clinical decision-making, medication changes, medication adherence, or broader engagement constructs. Third, probabilistic class assignment may introduce misclassification, particularly for intermediate phenotypes; although cross-resolution agreement supported overall robustness, the lower concordance of the Stable Low Observed Retention phenotype (62.1%) indicates sensitivity to temporal specification. Fourth, substantial administrative right censoring (79.2% with <24 months of follow-up) limits interpretation of trajectory shapes. Declining contact patterns among low-contact phenotypes may reflect limited observation windows rather than confirmed disengagement. Fifth, the cohort was derived from a single quality improvement program and may not represent the broader hypertensive population. Finally, the absence of linked cardiovascular outcomes also prevents assessment of whether observed differences in treatment documentation or BP control translate into differences in long-term cardiovascular risk.

### Future direction

Future studies should address three priorities. First, separating administrative censoring from true loss to follow-up is essential before clinical implementation. This may require complete observation-window designs, inverse-probability-of-censoring weighting, linkage to external healthcare data, or joint modelling of contact and censoring processes, followed by validation in independent, multi-site cohorts.[22] Second, linking observed contact trajectories with clinical outcomes is needed to establish prognostic relevance. A key hypothesis is that trajectory membership identified within the first 6 months after the index visit predicts 5-year major adverse cardiovascular events after adjustment for baseline BP and comorbidity burden. Another testable hypothesis is that treatment documentation gradients, rather than absolute BP differences, provide the stronger predictor of cardiovascular risk, consistent with the process-oriented interpretation of these findings. Third, prospective evaluation of early risk identification approaches combining baseline characteristics with early visit patterns may enable targeted outreach for patients at risk of declining observed contact. Pragmatic trials are needed to determine whether such interventions improve longitudinal contact, treatment documentation, BP control, and ultimately cardiovascular outcomes.[23, 24]

## CONCLUSION

This study identified four distinct patterns of observed EHR contact among patients with hypertension, reproducible across 1-month and 3-month interval models and supported by bootstrap validation (mean ARI 0.966–0.969; 100% convergence). These phenotypes ranged from rapid early decline in observed contact to sustained high contact over 36 months, demonstrating substantial heterogeneity in longitudinal visit patterns. After adjustment for demographic and clinical characteristics, higher-contact phenotypes consistently showed greater treatment documentation rates than the Low Observed Follow-up group, whereas associations with BP control were smaller and less consistent, suggesting that observed contact trajectories are more strongly related to care processes than to BP outcomes. Fixed time-point analyses further showed minimal between-group differences in absolute BP levels and indicated that apparent first-to-last BP improvements were influenced by differences in observation duration. Overall, latent class trajectory modelling of repeated binary visit indicators provides a useful framework for characterizing heterogeneity in EHR-based contact patterns. Future studies incorporating improved censoring adjustment, external validation, and linkage to cardiovascular outcomes are needed to determine whether these observed trajectories can support risk stratification and targeted interventions in hypertension care.

## CONFLICTS OF INTEREST

None.

## CONTRIBUTION STATEMENT

JY designed the study, contributed to the data analyses, and the writing of the manuscript. HC contributed to data analyses and the writing of the manuscript. All authors read and approved the final version of the manuscript.

## DATA AVAILABILITY

Datasets are available from the corresponding author on reasonable request and completion of appropriate data sharing agreements. The relevant code and analyses are available at https://github.com/hxcconnie-star/latent_class_trajectory_hypertension_phenotypes.

## ACKNOWLEDGMENTS

We thank the participating EvidenceNOW practices, clinicians and quality-improvement staff for their commitment to improving cardiovascular preventive care in their communities, and the patients whose deidentified data made this work possible.

## FUNDING

None.

## Supplementary

**Table S1.** Model selection for 1-month interval.

| K | Log-likelihood | BIC | AIC | Relative entropy | Min APP | Min OCC |
| --- | --- | --- | --- | --- | --- | --- |
| 1 | -277809.8 | 555986.5 | 555691.5 | - | - | - |
| 2 | -257753.9 | 516251.8 | 515653.7 | 0.847 | 0.925 | 8.68 |
| 3 | -254350 | 509821.3 | 508920.1 | 0.837 | 0.902 | 7.07 |
| 4 | -252291.3 | 506081 | 504876.6 | 0.821 | 0.77 | 6.49 |
| 5 | -251151.6 | 504178.7 | 502671.2 | 0.714 | 0.775 | 4.84 |
| 6 | -250450.3 | 503153.3 | 501342.7 | 0.741 | 0.742 | 5.46 |

**Table S2.** Model selection for 3-month interval.

| K | Log-likelihood | BIC | AIC | Relative entropy | Min APP | Min OCC |
| --- | --- | --- | --- | --- | --- | --- |
| 1 | -141829 | 283780 | 283681.7 | - | - | - |
| 2 | -126705 | 253664.8 | 253460 | 0.824 | 0.922 | 7.69 |
| 3 | -124893 | 250172.8 | 249861.4 | 0.769 | 0.782 | 5.86 |
| 4 | -123696 | 247911 | 247493.2 | 0.771 | 0.778 | 5.39 |
| 5 | -123397.6 | 247447.6 | 246923.2 | 0.700 | 0.677 | 4.66 |
| 6 | -123125.7 | 247036.3 | 246405.5 | 0.622 | 0.671 | 3.30 |

**Table S3.**
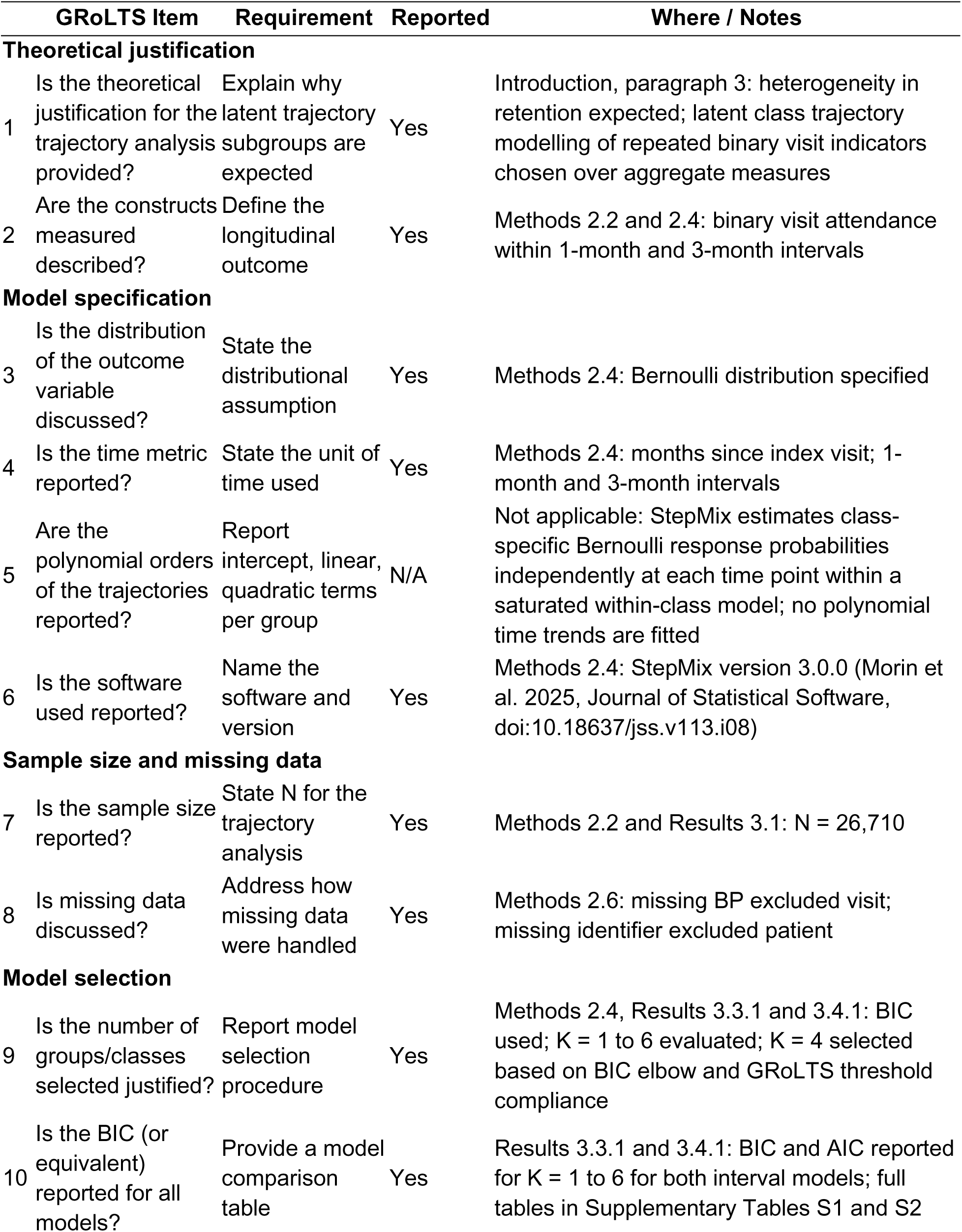
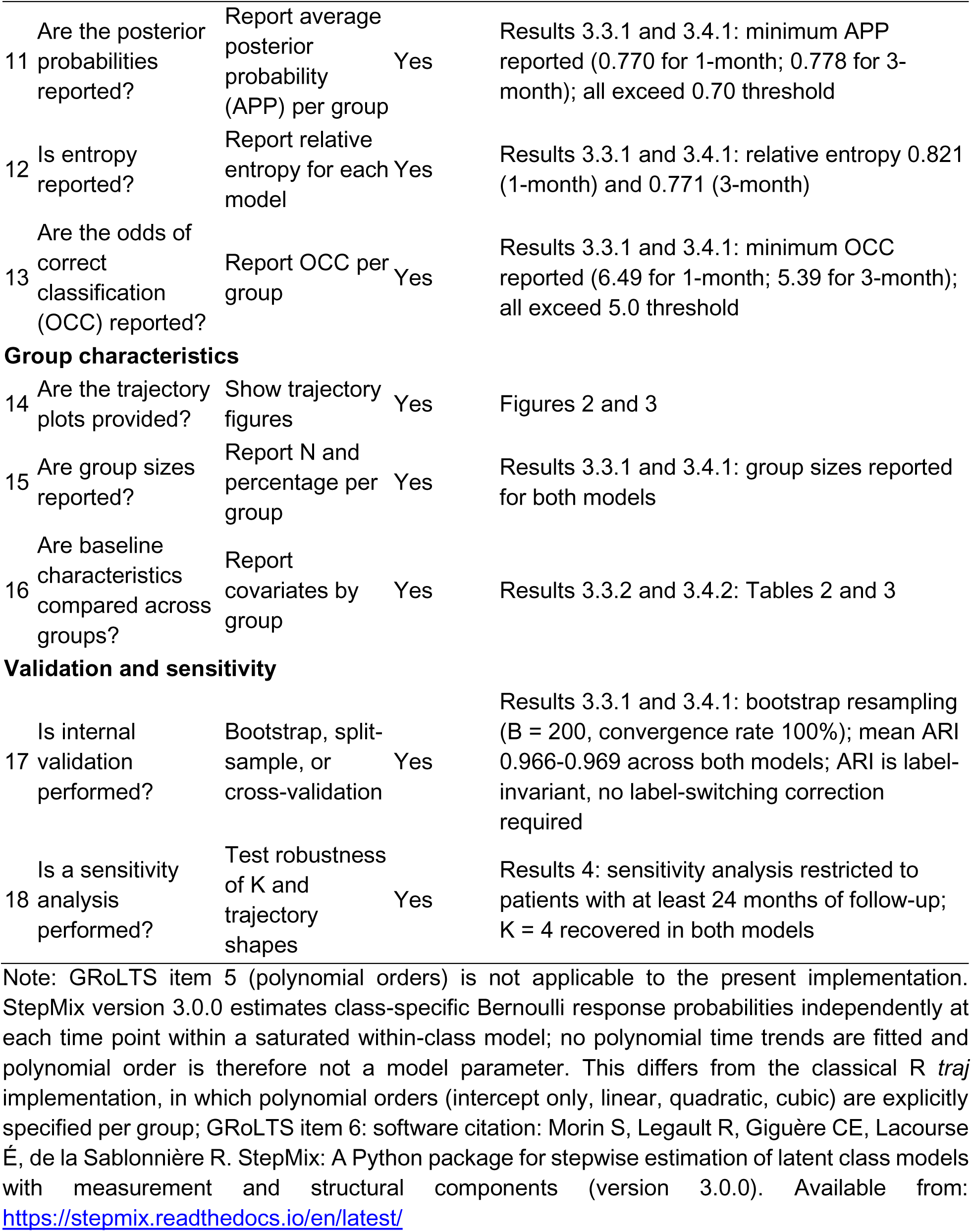
GRoLTS compliance table.

**Table S4.** Number of Patients with Available BP Measurement at Fixed Time Points by Trajectory Group (1-Month Interval)

| **Group** | **N total** | **N at 12 mo (%)** | **N at 24 mo (%)** | **N at 36 mo (%)** |
| --- | --- | --- | --- | --- |
| Low Observed Follow-up | 18,308 | 9,046 (49.4%) | 3,747 (20.5%) | 959 (5.2%) |
| Stable Low Observed Retention | 4,879 | 3,863 (79.2%) | 4,022 (82.4%) | 2,335 (47.9%) |
| Delayed Decline in Observed Visits | 2,945 | 2,703 (91.8%) | 1,318 (44.8%) | 373 (12.7%) |
| Sustained Observed Retention | 578 | 409 (70.8%) | 403 (69.7%) | 278 (48.1%) |

**Table S5.** Number of Patients with Available BP Measurement at Fixed Time Points by Trajectory Group (3-Month Interval)

| **Group** | **N total** | **N at 12 mo (%)** | **N at 24 mo (%)** | **N at 36 mo (%)** |
| --- | --- | --- | --- | --- |
| Low Observed Follow-up | 18,008 | 8,718 (48.4%) | 3,462 (19.2%) | 880 (4.9%) |
| Stable Low Observed Retention | 3,320 | 2,421 (72.9%) | 2,586 (77.9%) | 1,479 (44.5%) |
| Delayed Decline in Observed Visits | 3,892 | 3,637 (93.5%) | 2,203 (56.6%) | 627 (16.1%) |
| Sustained Observed Retention | 1,490 | 1,245 (83.6%) | 1,239 (83.2%) | 959 (64.4%) |

**Table S6.** Model selection for sensitivity analysis, 1-month interval.

| K | Log-likelihood | BIC | AIC | Relative entropy | Min APP | Min OCC |
| --- | --- | --- | --- | --- | --- | --- |
| 1 | -96874.8 | 194060.0 | 193821.6 | - | - | - |
| 2 | -93344.3 | 187318.2 | 186834.6 | 0.921 | 0.913 | 8.14 |
| 3 | -92549.4 | 186047.4 | 185318.7 | 0.720 | 0.835 | 4.69 |
| 4 | -92257.2 | 185782.2 | 184808.4 | 0.737 | 0.753 | 4.49 |
| 5 | -92004.1 | 185595.1 | 184376.2 | 0.755 | 0.758 | 6.05 |
| 6 | -91756.0 | 185418.1 | 183954.1 | 0.715 | 0.734 | 6.16 |

**Table S7.** Model selection for sensitivity analysis, 3-month interval.

| K | Log-likelihood | BIC | AIC | Relative entropy | Min APP | Min OCC |
| --- | --- | --- | --- | --- | --- | --- |
| 1 | -41903.4 | 83910.3 | 83830.8 | - | - | - |
| 2 | -40112.6 | 80440.9 | 80275.3 | 0.683 | 0.850 | 6.63 |
| 3 | -39595.0 | 79517.8 | 79266.1 | 0.679 | 0.814 | 10.87 |
| 4 | -39400.3 | 79240.5 | 78902.6 | 0.648 | 0.741 | 7.23 |
| 5 | -39155.8 | 78863.6 | 78439.6 | 0.679 | 0.752 | 11.89 |
| 6 | -39026.9 | 78717.9 | 78207.8 | 0.657 | 0.708 | 7.30 |

**Figure S1.**
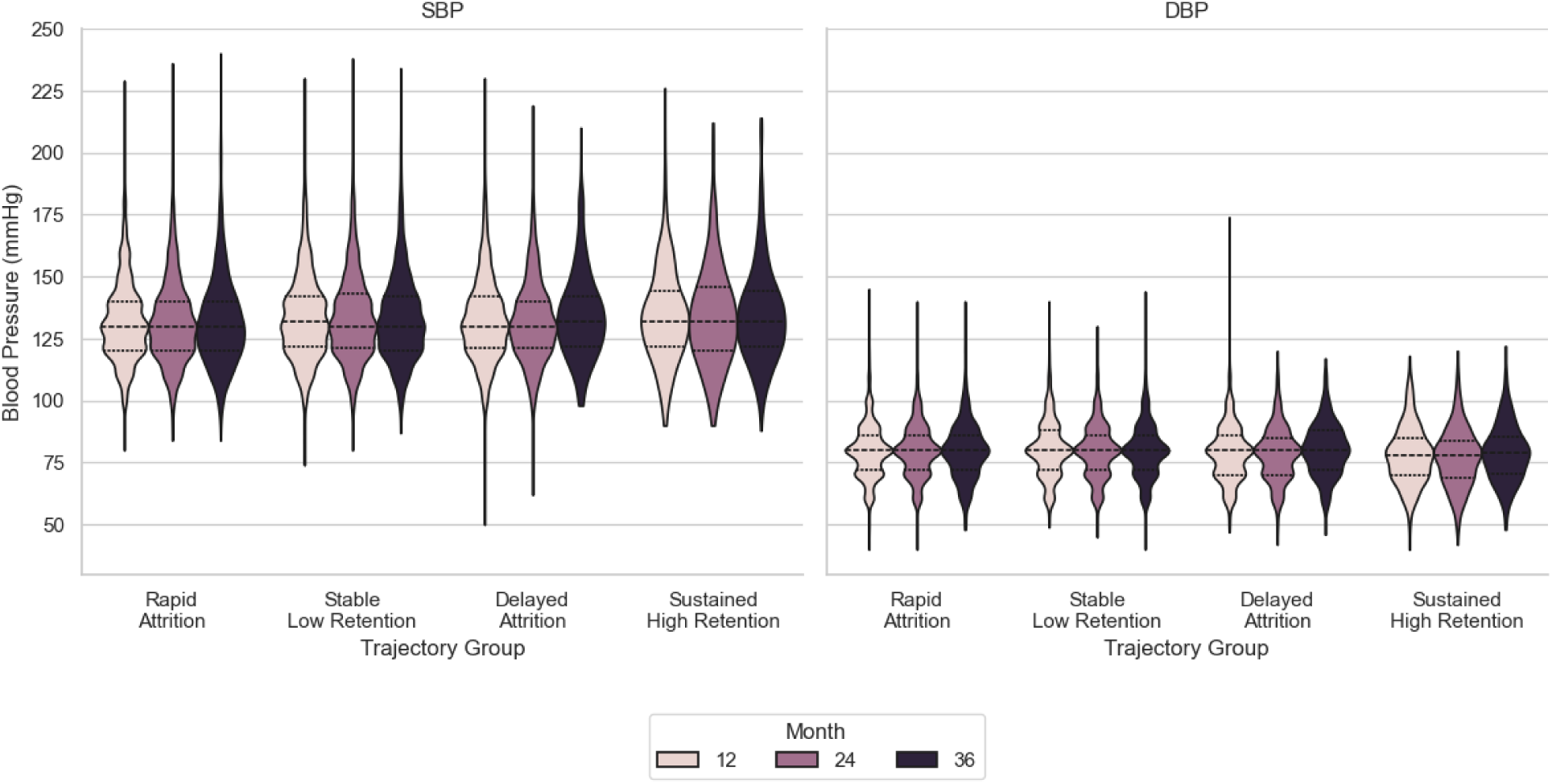
Time-point BP outcomes at 12, 24, 36 months (1-month)

**Figure S2.**
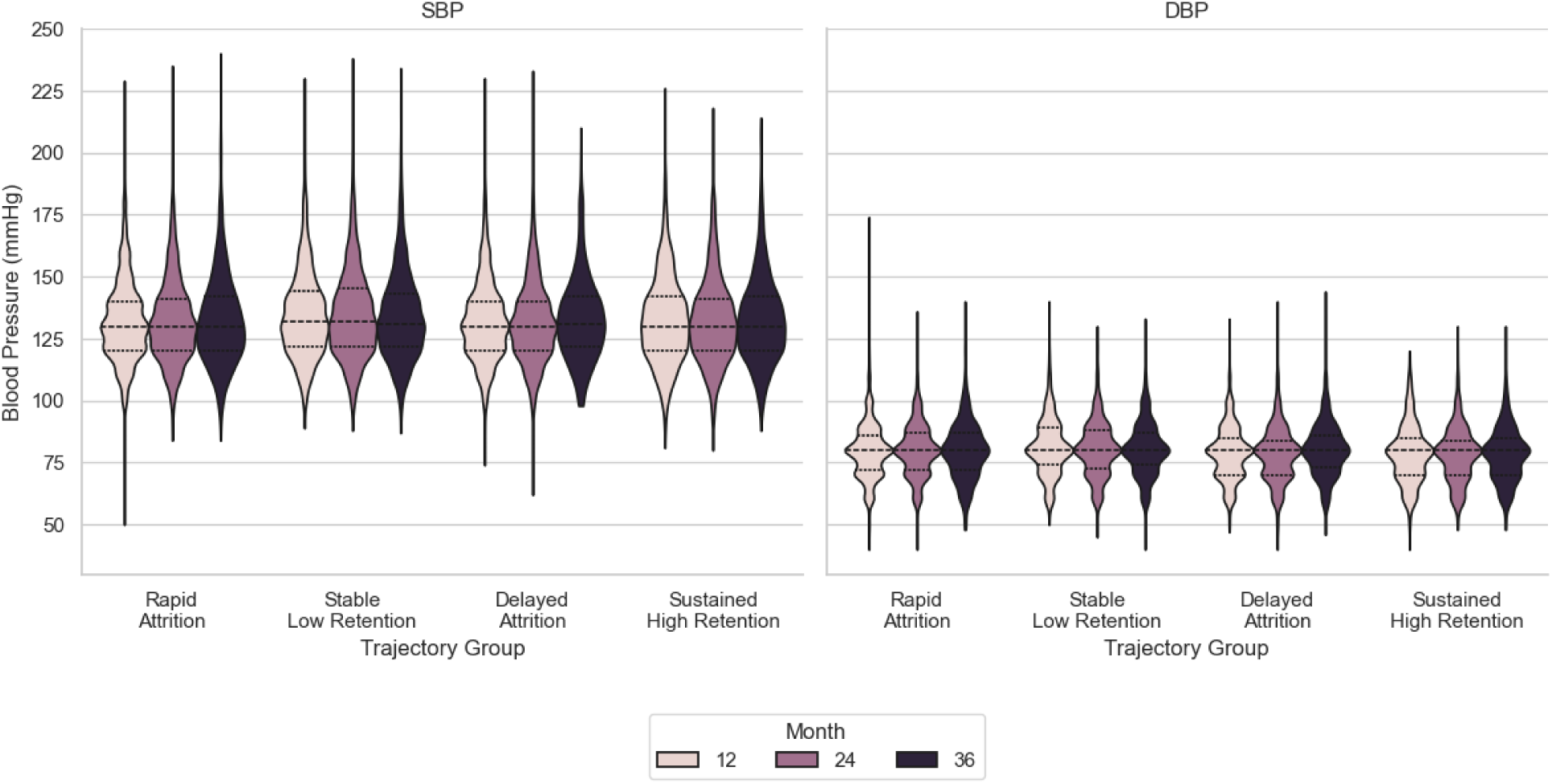
Time-point BP outcomes at 12, 24, 36 months (3-month)

**Figure S3.**
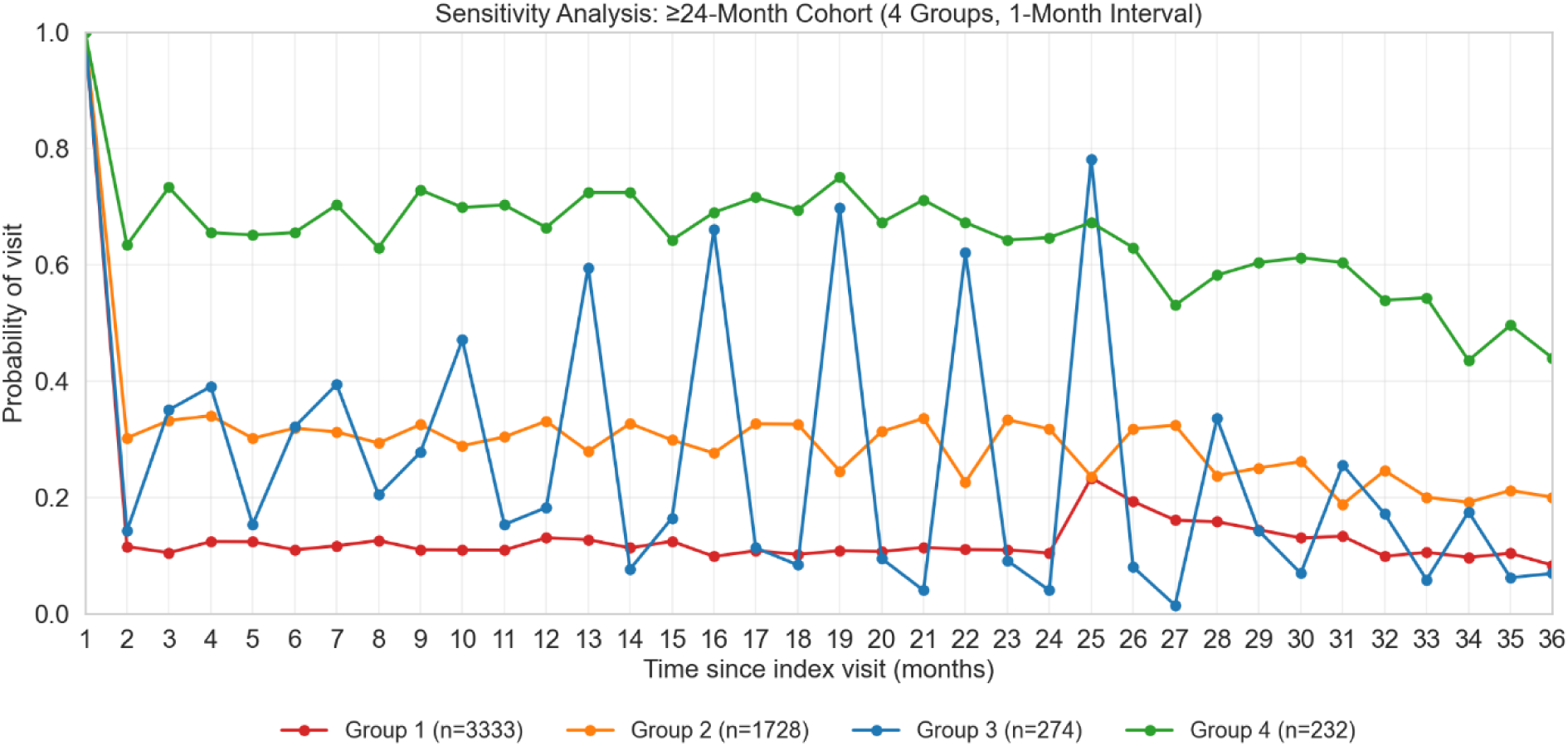
Trajectories by 1-month interval on patients with at least 24 months follow-up.

**Figure S4.**
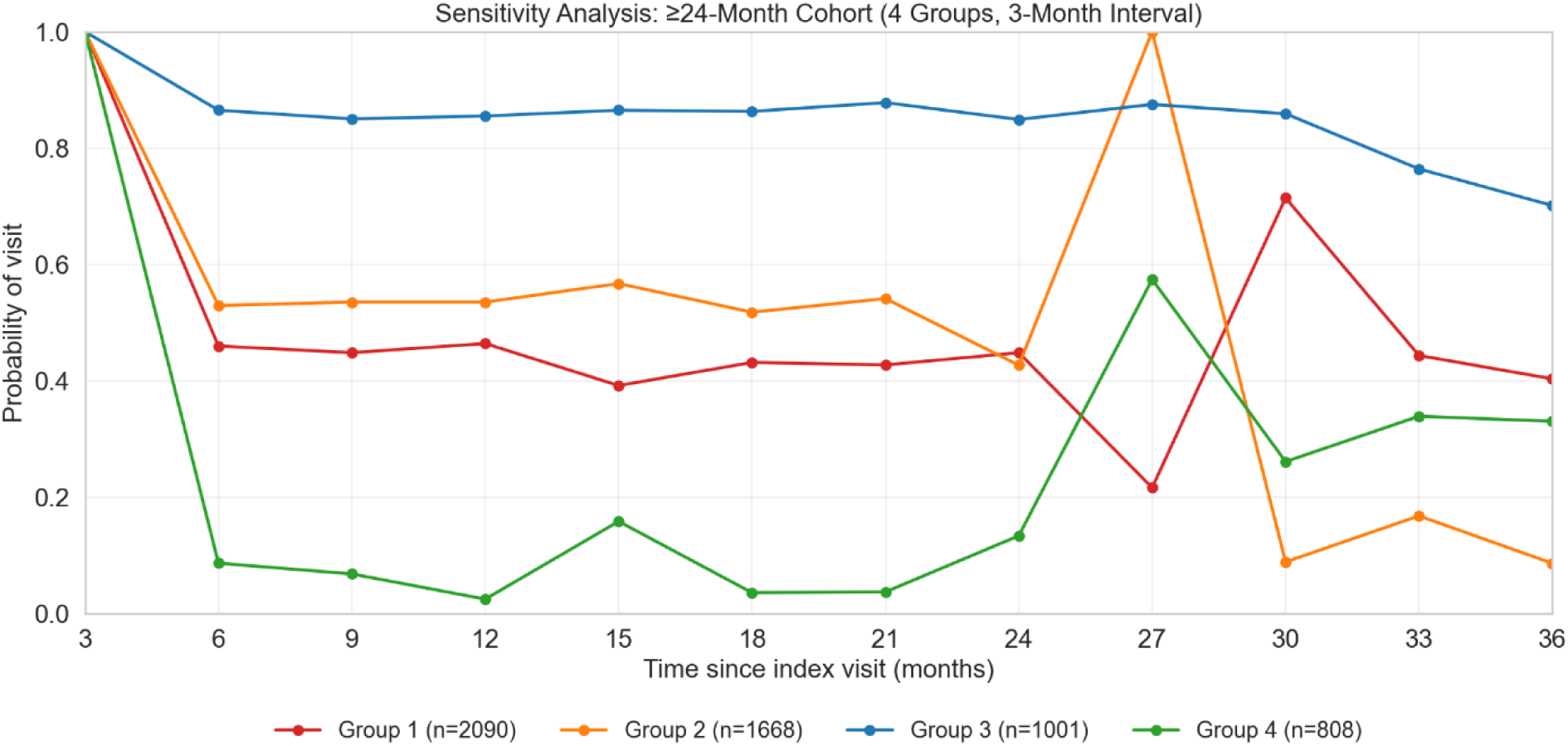
Trajectories by 3-month interval on patients with at least 24 months follow-up.

## Notes

### Competing Interest Statement

The authors have declared no competing interest.

### Author Declarations

This study utilized the dataset from a quality improvement program (EvidenceNOW). All the data were de-identified and the study was approved by Northwestern University's Institutional Review Board.

